# Beyond Symptoms: WHODAS as a Biopsychosocial Measure to Complement Functioning Assessment in Parkinson’s Disease

**DOI:** 10.1101/2025.07.31.25331974

**Authors:** Nathalia de Brito Pereira, Rebeka Amanda Dias, Katia Cirilo Costa Nobrega, Isaíra Almeida Pereira da Silva Nascimento, Rafaela Pires Bocicovar, Carolina Gonçalves Santana, Letícia Rocha Matos, Fernando Fachine da Silva Fidelis, Gustavo Thomazella Cheque de Campos, Luiza Mattos Aranha, Graziele Cardoso dos Santos, André Frazão Helene, Antonio Carlos Roque, Carsten Eggers, Maria Elisa Pimentel Piemonte

## Abstract

Functionality is considered the third health indicator, complementing the traditional mortality and morbidity metrics. However, functionality is rarely assessed systematically in Parkinson’s disease (PD) clinical practice and research, where symptom-based scales predominate. Identifying declines across various dimensions of functionality in PD is essential for patient education, disease management, and guiding new intervention strategies. The World Health Organization Disability Assessment Schedule 2.0 (WHODAS 2.0) is a comprehensive assessment tool developed by the World Health Organization (WHO) based on the International Classification of Functioning, Disability and Health (ICF) to standardize the measurement of functionality and disability impacts across diverse health conditions and cultural contexts. This study aimed to characterize functionality across PD severity stages using the WHODAS 2.0, independent of age, sex, socioeconomic status (SEC), and education. A total of 352 patients were divided into four clinically severity PD stage groups according to the Hoehn & Yahr (H&Y) scale. The age, sex, SEC and education levels were controlled to guarantee paired groups. All participants were evaluated remotely using the Telephone - Montreal Cognitive Assessment (T-MoCA), Beck Depression Inventory (BDI), Movement Disorder Society – Unified Parkinson’s Disease Rating Scale, Part I (MDS-UPDRS I) and Part II (MDS-UPDRS II) and WHODAS 2.0. The most affected functionality dimensions were Mobility, Activities of Daily Life related to the Household, and Participation. The nonparametric Kruskal-Wallis test revealed a significant effect of the group for all functionality dimensions. Notably, Mobility, Activities of Daily Life related to the Household, and Self-Care showed a gradual decline starting from stage 1 of H&Y. In contrast, Cognition, Getting Along and Participation exhibited progressive impairment only from stage 2 of H&Y. This study is the first to describe functionality in PD using WHODAS 2.0 across severity stages controlling for key demographic factors. Findings highlight that WHODAS captures functional limitations not identified by symptom-focused scales such as MDS-UPDRS. Incorporating WHODAS into routine assessment may improve patient-centered care by informing interventions targeting functional limitations from early disease stages.

**Highlights:** WHODAS 2.0 captures functionality domains not assessed by MDS-UPDRS.

Functional decline in mobility, self-care, and household tasks starts at early PD stages.

Participation, cognition, and interpersonal relationships decline from stage 2 onwards.

Functionality decline is independent of age, sex, education, and socioeconomic status.

WHODAS provides a biopsychosocial assessment essential for person-centered PD care.

## Introduction

Since 2001, the International Classification of Functioning, Disability and Health (ICF) developed by the World Health Organization (WHO) has been considered the global standard for defining health and health-related conditions (1). According to the ICF, functionality is influenced by a complex interplay of biological, psychological, and social factors and can be impacted by various health conditions, such as Parkinson’s disease (PD) (2). Functionality is considered the third critical health indicator, complementing the established mortality and morbidity indicators (3). According to this model, disability, i.e., decreased level of functionality, is conceptualized as a health experience that occurs in a context rather than as a problem that resides solely in the individual. This approach allows for a more comprehensive assessment of health conditions and considers the influences of individual, social, and environmental factors on functioning and disability (1).

PD has the highest disability weight according to the Global Burden of Disease (4). The PD progression is caused by the progressive loss of neurons in the brain and the peripheral autonomic nervous system. The clinical progression is multidimensional and includes the effects of aging (5). The progressive neurodegenerative process may begin several years before the diagnostic and affects motor function, with symptoms such as tremors, rigidity, bradykinesia, and postural instability and non-motor functions, with symptoms such as cognitive decline, depression, anxiety, and autonomic dysfunction (5). Together, these symptoms, associated with motor complications that may arise as a result of L-dopa therapy, contribute significantly to increased disability and reduced quality of life (QoL) quality in PD (6). Besides disease-related factors, lack in social support is also a significant factor that contribute for decreased QoL (7), and environment (7). Barriers contribute to increased limitations and restrictions in the social participation (2). Age (8), sex (9), socioeconomic condition (SEC) (10, 11), and education (12, 13,14) are other factors that can impact the onset, severity, and disease progression.

Motor deterioration and disability level ranged from 2.4 to 7.4% per year, and standard deviations indicated that there was considerable variability of progression rates between individuals. The motor impairment and disability level in PD progress show a large variability between the individuals, ranged from 2.4 to 7.4 per year (15). The annual rate decline in motor function in activities of daily living (ADL) are similar (16) and comparable with annual reduction in neurodegenerative process in striatum (17,18).

The Hoehn & Yahr (H&Y) scale has been the most commonly used scale worldwide to describe the severity of PD since its introduction (19). Higher H&Y stages are correlated with progressive dopaminergic loss (20), and the H&Y stages are strongly correlated with standardized scales of motor impairment, disability, and QoL (21). The severity of the disease is classified into five stages, where patients in stage 1 are only minimally disabled, while those in stage 5 experience severe disabilities (22). The progression from one stage to the next is clearly defined and not influenced by evaluator bias or temporary changes in motor function caused by medication (23). However, the transition time from one stage to another range widely between individuals, being inversely associated with age and level of motor impairment at the time of diagnosis (24)

The progressive disease severity decreases the ADL independence (25), the QoL (26), and increases the handicap (27). Limitations in ADL are indicative of disability level (28), disease progression (29), and the effectiveness of interventions (30, 31). Although ADL independence is a key aspect of assessing disability level, it is not the same as functionality, which is much more comprehensive (1). QoL has also been often used for the same functionality. Although conceptually, QoL and functionality share several constructs and are interchangeable, they are also not the same. QoL reflects subjective well-being (i.e., feeling satisfied with one’s performance in a specific life domain) (32), while functionality indicates objective performance in each life domain (33).

As the population ages and life expectancy increases, the number of people with PD for an extended period will increase, leading to a rise in the number of people experiencing high levels of disability (4). Rehabilitation stakeholders should adopt functionality as the primary outcome as a political priority worldwide (34). Then, knowing the progression of functionality and disability in PD is a key step for guiding health care for people who live with the disease. However, measuring functionality and disability presents challenges as they encompass various aspects of life and involve interactions between individuals and their environment. The World Health Organization Disability Assessment Schedule 2.0 (WHODAS 2.0) is a comprehensive assessment tool created by the WHO to measure the impact of a health condition on a person’s functionality and disability across different cultures (1). It was developed from the ICF framework and proven to be a valid, reliable, and efficient self-reported tool for measuring disability (35) short- and long-term effects of interventions (36, 37) and long-term functionality trajectory (38). The tool’s cross-cultural applicability (39), reliability, validity (40), and usefulness in health services research (41) have been confirmed through systematic field studies.

Although other generic instruments for assessing health conditions can be mapped to ICF, they do not clearly distinguish between measuring symptoms, disability, and subjective appraisal (40). WHODAS 2.0 provides a standard metric for the impact of any health condition in terms of functioning. It also assesses disability in a culturally sensitive way across a standard rating scale (41). Although there are several PD-specific tools able to measure disease progression, such as the MDS-UPDRS, WHODAS 2.0 can provide a more comprehensive perspective on the disease’s progression by incorporating domains related to social participation, autonomy, and work-related activities, which are key aspects for understanding the real-life challenges faced by people with PD. A recent review of the WHODAS 2.0 application among elderly populations confirmed its validity in measuring the disability levels across several health conditions, including PD, and recommended its global utilization in medical and rehabilitation contexts since it provides a uniform evaluation of functionality and performance (42).

Several studies have shown that WHODAS 2.0 is effective in assessing the functional impact of PD. However, there are significant gaps in understanding how disability progresses at different stages of the disease. While previous research has examined WHODAS 2.0’s psychometric properties (43), the relationship between cognitive impairment and functional disability (44), and specific functional deficits in early-onset versus late-onset PD (45), these studies have not systematically analyzed the decline in functionality across stages of disease severity. Other previous studies have focused on moderate-to-advanced PD (46, 47), failing to capture early functional impairments that could be critical for timely interventions. Additionally, previous studies have predominantly used a single scoring method for WHODAS 2.0, limiting the scope of functional assessment.

Another significant limitation in the current evidence is the lack of comprehensive sociodemographic control in evaluating functional decline. Factors such as age (8, 45), sex (9, 48–50), socioeconomic status (SES) (10, 11), and education (51) can influence disability progression, yet their impact has not been adequately accounted for in existing studies. Moreover, while recent advancements in machine learning have enabled the development of a shorter, efficient WHODAS-based assessment (52), these models have not examined longitudinal functional decline nor compared disability trajectories across H&Y stages.

To address these gaps, the present study aims to investigate the progressive decline in functionality across different PD severity stages using WHODAS 2.0 while controlling sociodemographic variables. By employing multiple scoring methods, this study seeks to provide a comprehensive and nuanced assessment of disability progression, identifying the earliest functional domains affected by PD. We hypothesize that WHODAS 2.0 will demonstrate a stage-dependent functional decline. Additionally, we anticipate that the impact of PD on functionality will remain significant even after controlling sociodemographic factors, reinforcing the robustness of WHODAS 2.0 as a standardized tool for assessing functional disability in PD, offering new evidence with important implications for patient care and disease management.

## Results

### Demographic and clinical features

A total of 352 participants completed the interview (Figure 1). Table 1 presents the demographic and clinical characteristics of the study participants according to H&Y classification. Most of the participants were male (55%) and identified as white (59%), while others identified as black (21%), multiracial (12%), or Asian (9%). As expected, there were no significant differences in age, race, sex, SEC, and education levels between the groups. However, as also expected, there were significant differences in levodopa equivalent daily dosage (LEDD), Telephone – Montreal Cognitive Assessment (T-MoCA), Inventory Depression Beck (BDI), and Movement Disorders Society – Unified Parkinson’s Disease Rating Scale, Part I (MDS-UPDRS I) and Part II (MDS-UPDRS II) scores (Table 1).

**Figure 1:**
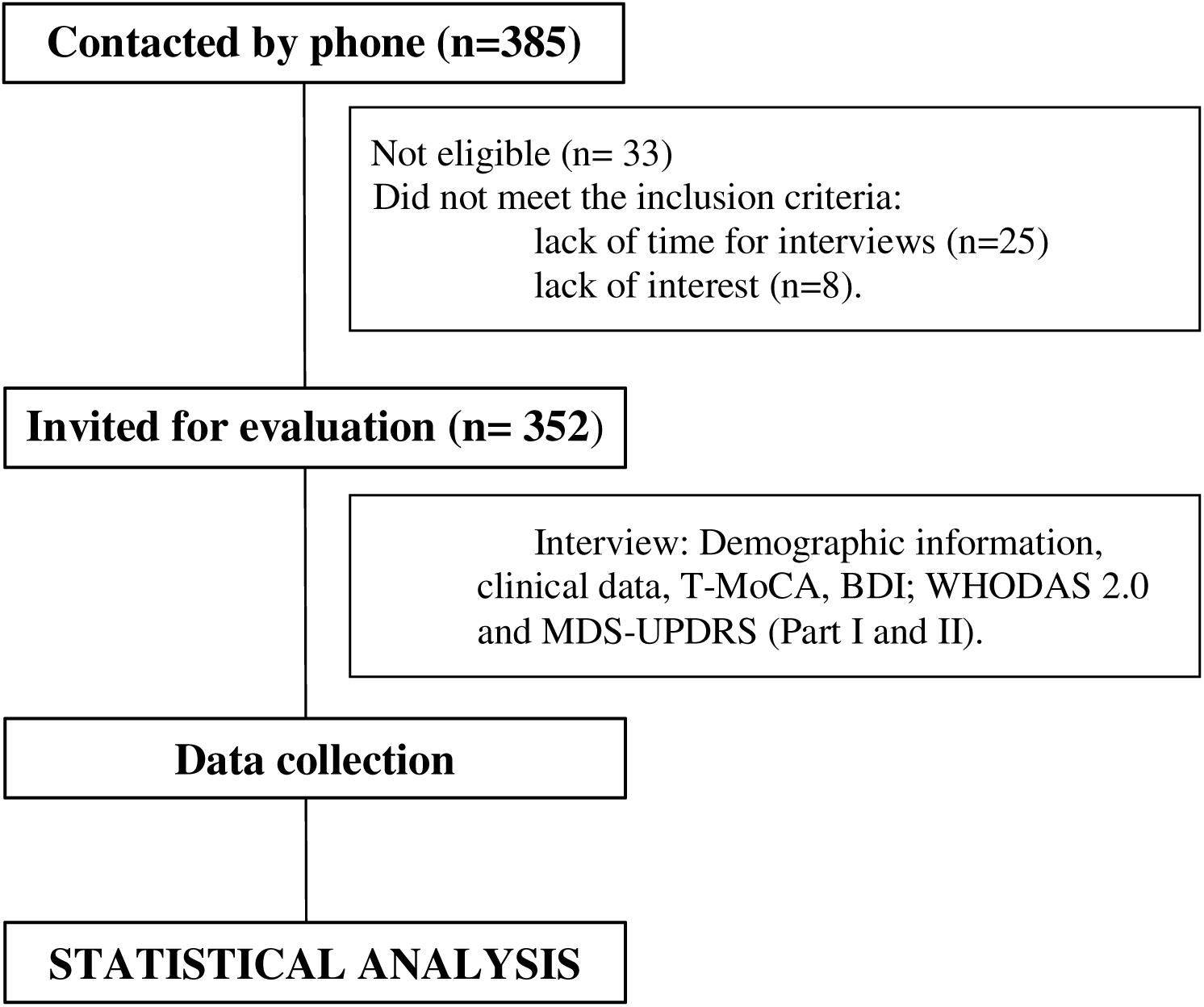
The schematic study designs. T-MoCA, Telephone – Montreal Cognitive Assessment; BDI, Inventory Depression Beck; WHODAS 2.0, World Health Organization Disability Assessment Schedule 2.0; MDS-UPDRS, Movement Disorders Society – Unified Parkinson’s Disease Rating Scale (Part I and II).

**Table 1:**
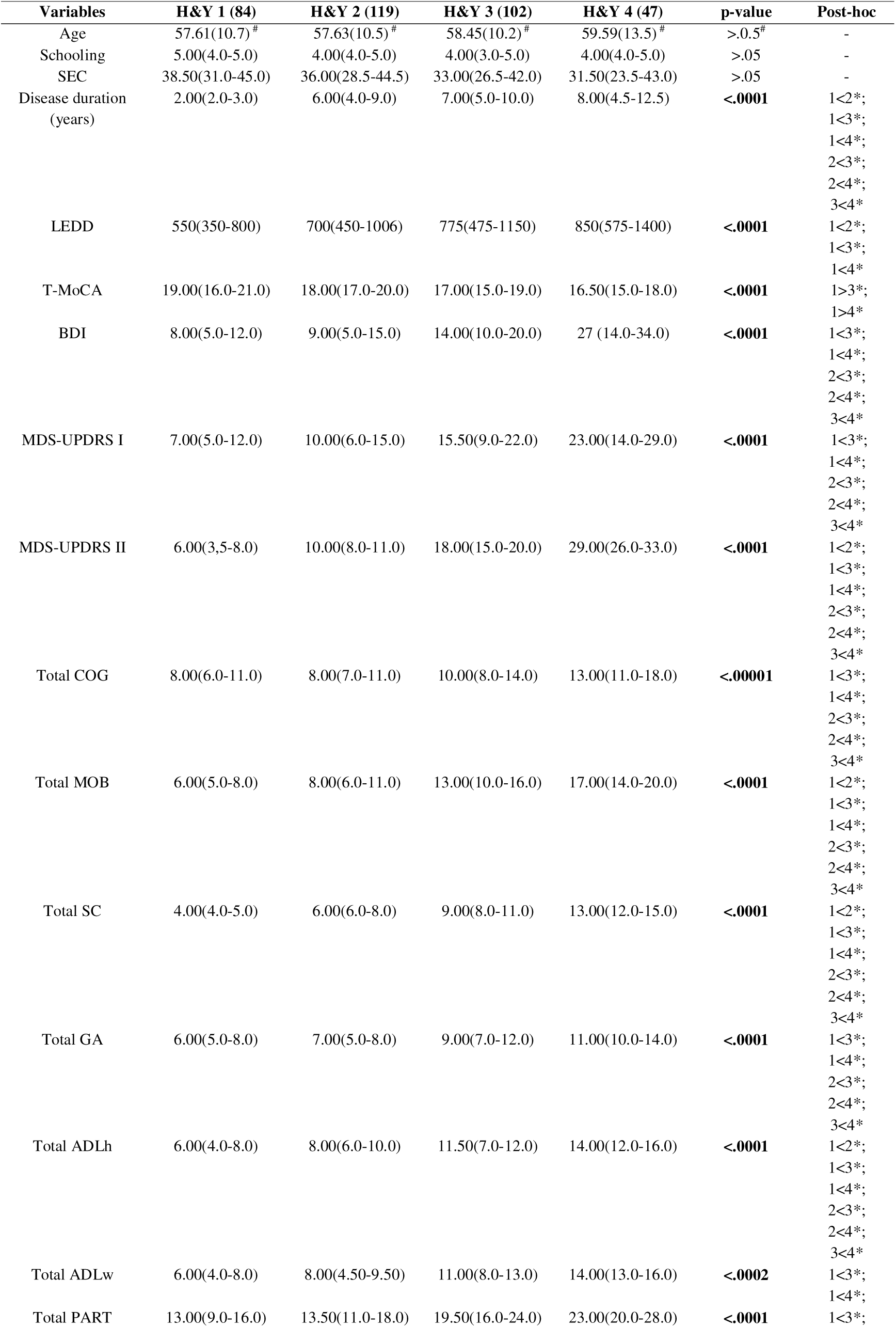
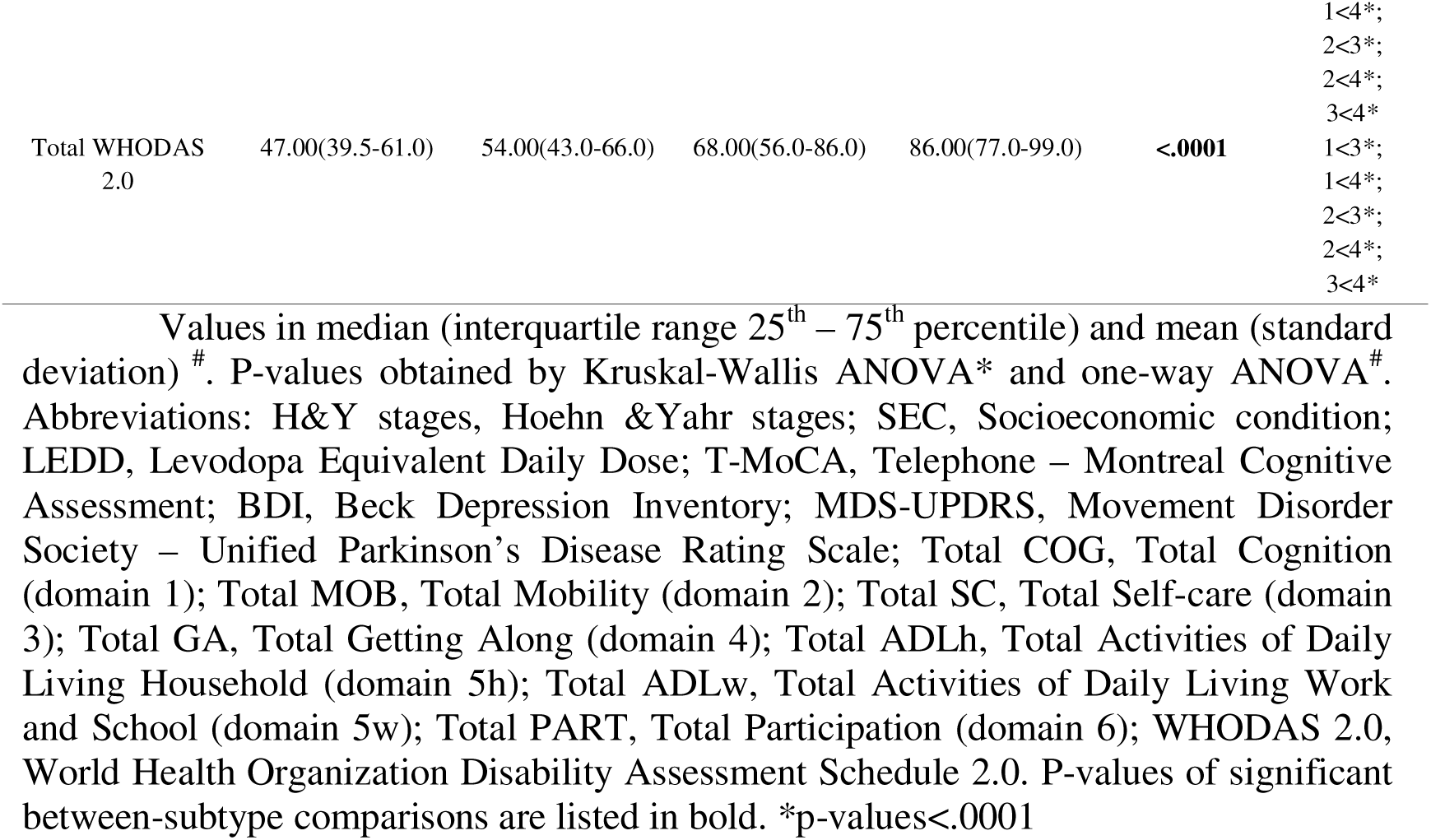
Demographic and clinical characteristics of the participants according to disease stages (H&Y)

### WHODAS 2.0

All participants were able to answer WHODAS 2.0. However, 32 participants had missing data for the self-care domain, specifically related to the ability to be alone, as they had not had the opportunity to be alone in the last 30 days. Additionally, in the Getting Along domain, 45 participants had missing data because they reported no sexual activity in the last 30 days. No ceiling or floor effects were noted. There was a wide distribution across disability levels on each dimension, indicating that WHODAS 2.0 effectively differentiated between participants.

According to the complex scoring system, the level of disability ranges from 1.88 (low) to 93.02 (high). According to dichotomy scoring, the activities with the most reported difficulties were completing household tasks quickly (87.3% of participants), getting dressed (74.2% of participants), and standing up from sitting (71.4% of participants). Additionally, 77.1% of participants reported difficulties related to the time spent managing their health condition, and 72.8% reported being emotionally affected by their health condition (Figure 2, Supplementary Table 1). Only 21% of participants reported keeping working, and more than 55% reported earning less money because of PD. Among the participants who reported being working, 46.67% were in stage 1, 32% in stage 2, 17.33% in stage 3, and only 4% in stage 4 (Supplementary Table 3-6).

**Figure 2:**
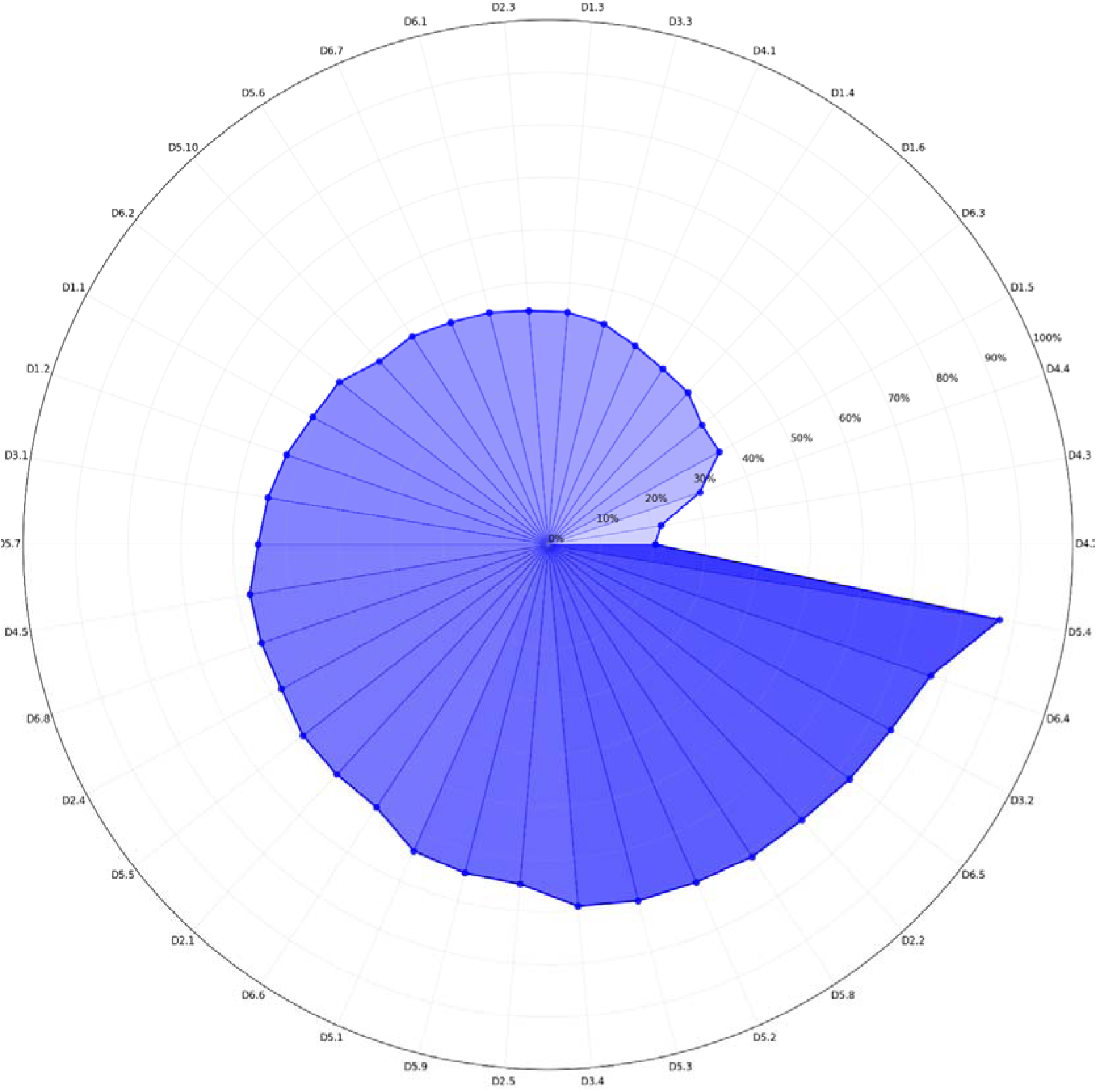
Percentage of reported disability for each WHODAS 2.0 question. This radar chart illustrates the distribution of “yes” responses for each question within the WHODAS 2.0 (World Health Organization Disability Assessment Schedule 2.0) domains with a maximum from 0% to 100% in 10% increments. The blue area shows the percentage of “yes” across all questions. The points on the chart indicate exact percentage values for each question. **D4.2:** Maintaining a friendship? *(20.4%);* **D4.3:** Getting along with people who are close to you? *(21.8%);* **D4.4:** Making new friends? *(30.6%);* **D1.5:** Generally understanding what people say? *(37.1%);* **D6.3:** How much of a problem did you have living with dignity because of the attitudes and actions of others? *(37.1%);* **D1.6:** Starting and maintaining a conversation? *(39.4%);* **D1.4:** Learning a new task, for example, learning how to get to a new place? *(39.9%);* **D4.1:** Dealing with people you do not know? *(41.4%);* **D3.3:** Eating? *(43.3%);* **D1.3:** Analyzing and finding solutions to problems in day-to-day life? *(44.5%);* **D2.3:** Moving around inside your home? *(44.8%);* **D6.1:** How much of a problem did you have joining in community activities (for example, festivities, religious or other activities) in the same way as anyone else can? *(45.6%);* **D6.7:** How much of a problem did your family have because of your health problems? *(46.2%);* **D5.6:** Doing your most important work/school tasks well? *(47.4%);* **D5.10:** Did you earn less money as the result of a health condition? *(47.4%);* **D6.2:** How much of a problem did you have because of barriers or hindrances in the world around you? *(50.4%);* **D1.1:** Concentrating on doing something for ten minutes? *(51.0%);* **D1.2:** Remembering to do important things? *(52.7%);* **D3.1:** Washing your whole body? *(54.1%);* **D5.7:** Getting all the work done that you need to do? *(55.3%);* **D4.5:** Sexual activities? *(57.6%);* **D6.8:** How much of a problem did you have in doing things by yourself for relaxation or pleasure? *(57.8%);* **D2.4:** Getting out of your home? *(58.7%);* **D5.5:** Your day-to-day work/school? *(59.2%);* **D2.1:** Standing for long periods such as 30 minutes? *(59.5%);* **D6.6:** How much has your health been a drain on the financial resources of you or your family? *(59.8%);* **D5.1:** Taking care of your household responsibilities? *(63.7%);* **D5.9:** Have you had to work at a lower level because of a health condition? *(64.6%);* **D2.5:** Walking a long distance such as a kilometer [or equivalent]? *(64.9%);* **D3.4:** Staying by yourself for a few days? *(69.1%);* **D5.3:** Getting all the household work done that you needed to do? *(70.0%);* **D5.2:** Doing your most important household tasks well? *(70.3%);* **D5.8:** Getting your work done as quickly as needed? *(71.1%);* **D2.2:** Standing up from sitting down? *(71.4%);* **D6.5:** How much have you been emotionally affected by your health condition? *(72.8%);* **D3.2:** Getting dressed? *(74.2%);* **D6.4:** How much time did you spend on your health condition or its consequences? *(77.1%);* **D5.4:** Getting your household work done as quickly as needed? *(87.3%)*.

Comparing the domains, the most affected were Activities Daily Living related to the Household, followed by Mobility and Participation (Figure 3, Supplementary Table 2).

**Figure 3:**
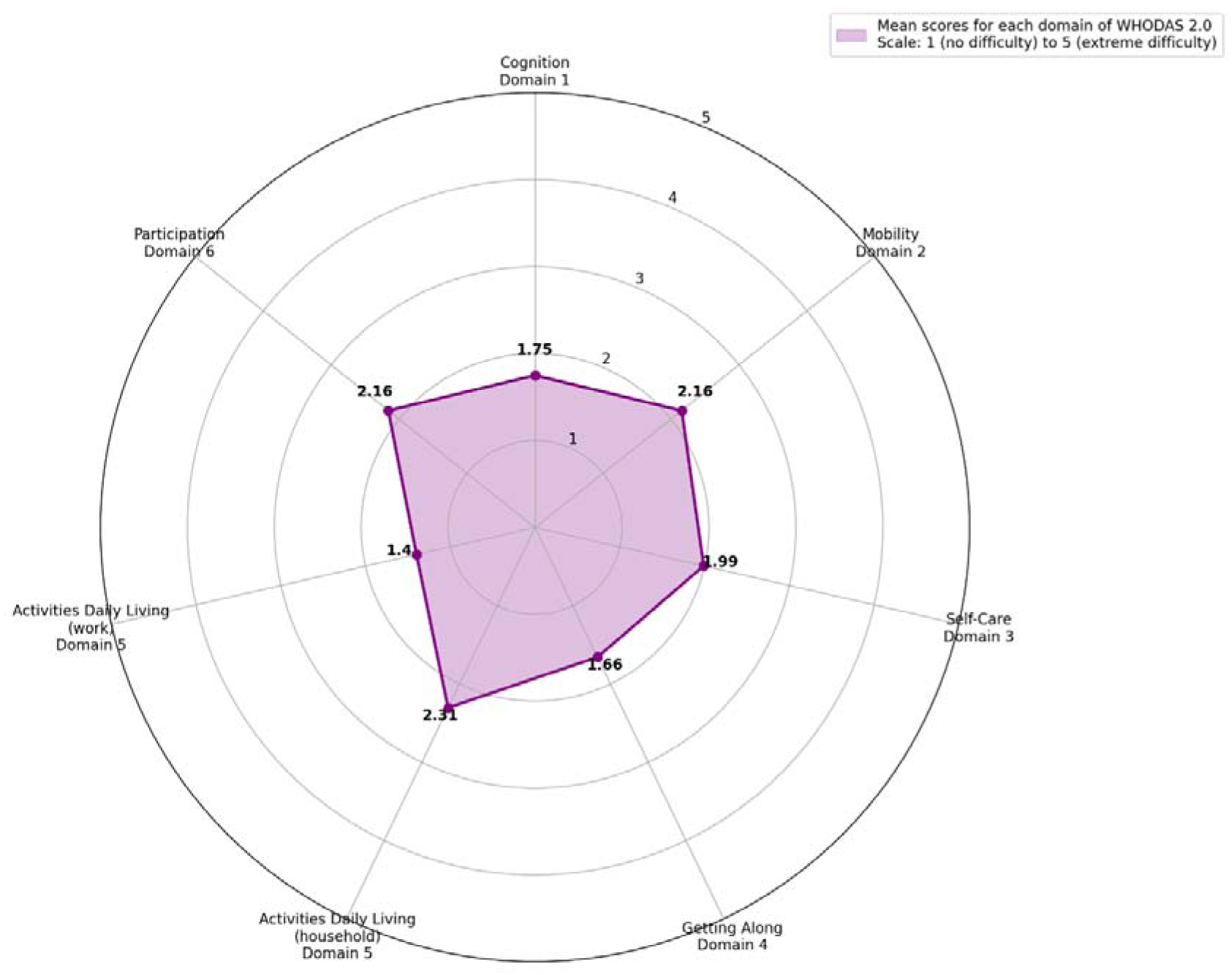
Comparison of WHODAS domains. The radar chart illustrates the average scores across different domains of the WHODAS 2.0 (World Health Organization Disability Assessment Schedule 2.0) questionnaire, with a maximum score of 5. Each axis represents a domain of functionality, and the purple line indicates the average score for each domain. Higher scores, closer to 5, indicate higher disability level in that domain. The seven domains represented are: 1- Cognition, 2 - Mobility, 3 - Self-care, 4 - Getting Along, 5h - Activities Daily Living Household, 5w - Activities Daily Living Work, 6 - Participation. The purpose of this figure is to provide a visual analysis comparing the most and least affected domains.

According to both scoring (simple and complex), the H&Y stages had a statistically significant impact on the WHODAS 2.0 total score and subscores from all domains. For the Mobility, Self-Care, and Activities Daily Living domains, the post-hoc test confirmed a significant difference between all H&Y stages (Table 1, Figure 4). However, for the total scores, Cognition, Getting Along, and Participation domains, the post-hoc test confirmed a significant difference between all stages, except between H&Y stages 1 and 2. For Activities Daily Living Work, significant differences were found from 1 to 3 and 4 disease stages.

**Figure 4:**
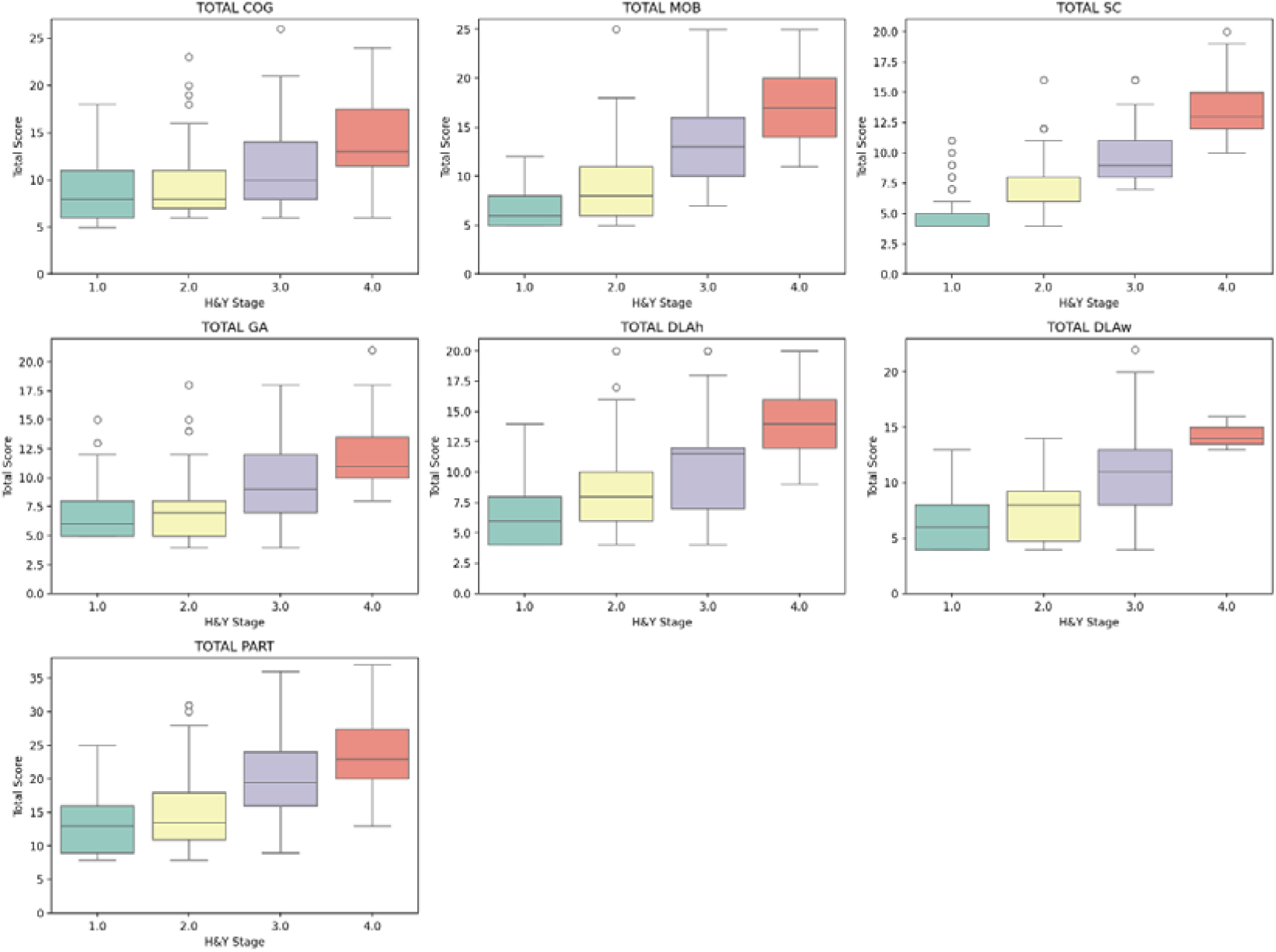
Comparison of H&Y stage for each WHODAS domains. Box represents the interquartile range (IQR, 25^th^ – 75^th^ percentiles), horizontal line inside represents median; whiskers extend to the most extreme data points within 1.5 times the IQR, Points represents individual outliers beyond the whiskers. Abbreviations: WHODAS, World Health Organization Assessment Schedule 2.0, Total COG, Total Cognition; Total MOB, Total Mobility; Total SC, Total Self-care; Total GA, Total Getting along; Total DLAh, Total Daily Life Activities Household; Total DLAw, Total Daily Life Activities Work/school; Total PART, Total Participation.

Additionally, a statistically positive high correlation was observed between WHODAS 2.0 total scores and MDS-UPDRS part I (R=.67, p-value<.00001) and MDS-UPDRS part II (R=.68, p-value<.00001), and moderate negative correlation between WHODAS 2.0 Cognition and T-MoCA (R=.33, p-value<.0001).

## Discussion

To our knowledge, this is the first study to systematically characterize the decline in each of the six functionality domains defined by the ICF framework across PD severity stages while controlling for age, sex, SEC, and education level. According to H&Y stages, disease severity affected the total WHODAS 2.0 scores and all sub-scores. The results show that Mobility, Self-Care, and Activities Daily Living Household domains are affected progressively from stage 1, while Cognition and Getting Along domains are progressively impacted from stage 2 of H&Y. These findings reveal that disease progression affects not only ADLs, commonly assessed by scales such as the MDS-UPDRS, but also other key dimensions of functionality, including participation, work-related activities, and interpersonal relationships.

Previous studies that utilized the WHODAS 2.0 to assess disability levels did not include patients in stage 1 of H&Y, did not use several scoring methods, and did not account for several confounding factors (43, 44, 46, 47). Therefore, our findings represent the first to demonstrate progressive functional decline assessed by WHODAS 2.0 from stages 1 to 4 of H&Y by three scoring methods, regardless of age, sex, socioeconomic status (SEC), and education level differences.

### WHODAS 2.0 domains progressively affected from stage 1 of H&Y

The Mobility domain, which assesses activities such as standing, moving around inside the home, exiting the home, and walking long distances, was progressively impaired from stage 1 of H&Y. Serrano-Duenas et al. found that Mobility was the most affected domain in PD. However, they did not include patients in stage 1 of the disease severity (43). Mobility deteriorates with disease progression (53). Gait and balance impairments are recognized as important symptoms in the later stages of PD (54). The transition from H&Y stage 2 to 3 marks a pivotal milestone in PD, when gait and balance impairment results in disability in many gait-dependent activities (55). Previous studies have shown a progressive decrease in community walking from stages 1 to 3 of H&Y (56). However, our results indicate that difficulties with Mobility can arise early in the disease, impacting the ability to move both indoors and outdoors. Nearly half of the participants in stage H&Y 1 reported difficulty walking long distances, with this difficulty worsening over time to affect over 95% of participants in stage 4 (Supplementary Table 3-6).

The Self-Care, which assesses independence for hygiene, dressing, eating, and staying alone, and Activities Daily Living Household, which assesses difficulty with day-to-day activities associated with domestic responsibilities, were also progressively affected from stage 1 of H&Y. While our findings indicate that Activities Daily Living Household is the most affected domain, it was previously identified as the third most affected WHODAS domain in a study that only included patients in stages 2-4 of H&Y (43). The inclusion of patients in stage 1 of H&Y may explain these contrasting findings. Previous studies have shown the association between disease severity and an increase in ADL dependence. Decreased independence in ADL is considered a feature of intermediate to advanced disease stages (25). The MDS-UPDRS II considers the level of dependency to perform ADL as a key issue to score disease severity (27). However, the results from WHODAS 2.0 expand our understanding of ADL disability by including Activities Daily Living Household in addition to Self-Care activities. We found that, besides walking, standing up from a sitting position, and getting dressed, which are part of the core physical actions most affected by PD (56), the most prevalent complaint was related to slowness in doing household tasks, reported by most participants even in stage 1 of H&Y (Supplementary Table 1). This slowness is probably associated with bradykinesia, a cardinal symptom of PD present from its onset (57).

Furthermore, the present results showed a rapid increase in difficulty in staying by oneself for a few days, from 11% in H&Y stage 1 to 78% in stage 2 and reaching severe or total disability for almost 95% of participants in stage 4 of H&Y (Supplementary Table 3, 4, 6). These results indicate a progressive level of dependence and the need for caregiver support (58, 59), which poses a high burden for both the family (60) and society (61). Therefore, preventing the loss of independence should be a therapeutic goal for interprofessional teams from the early stages of the disease.

The level of disability in Activities Daily Living Work increases significantly only from stages 1 to 3 and 4 of H&Y. This result should be interpreted with caution, as there was a high percentage of missing data in this domain associated with a high proportion of patients who were neither working nor studying. The number of participants who continued working declined drastically from stages 1 to 3 and 4 of H&Y. Although age could be a contributing factor to these results (45), no age differences were observed between the H&Y stages in this study. This drastic reduction should probably be associated with increased impairment of mobility and cognition (62). Further studies are needed to investigate the factors that can explain the difficulties in maintaining Activities Daily Living Work.

### WHODAS 2.0 domains progressively affected from stage 2 of H&Y

The Cognition domain, which includes concentrating, remembering, problem-solving, learning, and communicating impairments, was progressively impaired from stage 2 of H&Y. In a previous study, Cognition was identified as the least affected WHODAS domains (41). However, in the current study, over 50% of participants reported experiencing difficulties in remembering important information and maintaining concentration for more than 10 minutes. While cognitive decline is often linked to aging (63), age alone cannot explain the findings observed in this study.

It is worth highlighting the comparison between the results of Cognition and T-MoCA. Although they were correlated, Cognition was able to identify progressive cognitive decline from stage 2 of H&Y, while T-MoCA could identify cognitive decline only from stages 1 to 3 and 4. These findings suggest that people with PD can perceive subtle changes in their cognitive capacity that objective tests like T-MoCA may not capture.

Cognitive decline may also be associated with the increased disability in the Getting Along domain, which assesses interactions with other people, including close connections (e.g., spouse or partner, family members, close friends) and strangers. The Getting Along progressive disability was also observed from stage 2. Although communication ability depends on cognition (64), voice alterations associated with PD can also contribute to difficulties interacting, particularly with strangers (65). Additionally, over half of the participants reported experiencing sexual difficulties, which were also evaluated within the Getting Along domain. Previous studies have shown that sexual dysfunction is a non-motor symptom that may present at the onset of the disease (66), increases progressively (67), and can be influenced by various multidimensional factors (68).

The decreased Cognition and Getting Along associated with Mobility disability may partly explain the reduction in Participation (69), which was also found from stage 2 of H&Y. PART assesses social dimensions, such as community activities, physical and social barriers to participation, and disease-related emotional and financial losses. Social integration has been regarded as the dimension of disability least affected by PD (46), but in our study, the Participation domain was the second most impacted, confirming other previous findings (43). Although a significant decrease in participation was only noticeable from stage 2, around 30% of participants in H&Y 1 reported experiencing difficulty in engaging with community activities and overcoming barriers in their environment (Supplementary Table 3). Furthermore, most of the participants reported experiencing emotional distress due to their health condition, with approximately 30% reporting social and familial problems related to the disease. Notably, more than half of the participants reported reduced income and a financial burden on themselves or their families due to PD. The progressive increase in the depressive symptoms shown by BDI confirms the emotional impact of disease severity.

Reduced mobility significantly limits participation in social activities (70), which in turn contributes to physical inactivity (71). Additionally, loneliness is a risk factor for cognitive decline (72). The combination of these impairments creates a reinforcing cycle: as both mobility and cognitive function decline, opportunities for social engagement decrease, leading to further reductions in physical activity and increased cognitive impairments (73).

Furthermore, although previous studies on suicide in PD have not explored the connection between reduced social participation and suicidal risk (74), a lack of participation and feelings of loneliness have been associated with an increased risk of suicide in individuals with physical disability (73). Thus, early multi-professional interventions should be implemented from the time of PD diagnosis to prevent mobility, cognitive, and communication impairments and their impact on the participation and social interaction of the patients. Psychological and social support should also be provided to help patients navigate the emotional and social challenges associated with the disease and maintain work activities and social interactions.

The correlation between WHODAS 2.0 and MDS-UPDRS parts I and II also merits comment. The MDS-UPDRS has been considered a gold standard for assessing PD severity (29, 45). Parts I and II have shown sensitivity for disease severity evolution (75,76) and QoL (77, 78). While the MDS-UPDRS has been widely accepted as the gold standard for assessing PD severity, it primarily focuses on motor and non-motor symptoms, failing to comprehensively capture the multidimensional impact of the disease on daily functioning. WHODAS 2.0, based on ICF, provides a more holistic evaluation of disability by encompassing domains such as mobility, self-care, participation, and work activities. Unlike the MDS-UPDRS, which is disease-specific, WHODAS 2.0 enables comparisons across various health conditions, facilitating a broader understanding of the functional decline associated with PD.

### Strengths and limitations

The present study has both strengths and limitations. The main strength was the recruitment process, which enabled the selection of participants from diverse geographical areas while maintaining consistency in age, sex, education level, and SEC. Significant relationships have been observed between disability according to WHODAS 2.0 and several socio-demographic factors such as sex, age, living status, need for help, and level of education (79, 80). These factors may also interfere in PD impact on functionality. Older patients had significantly higher scores in motor and nonmotor symptoms and handicap (27). Sex is a key role in the features of PD and may influence the mechanisms involved in disease development (9, 48–50). SEC (10, 11) and education level (51) were associated with a higher chance of PD (81), mortality (11), non-motor symptoms severity (81), and ambulatory capacity (54). Thus, the absence of significant differences between the groups in this study has certainly reduced the impact of these secondary factors on functionality, there by allowing the analysis of the disease severity effect alone.

Other strengths were the inclusion of participants in stage 4 of H&Y, addressing the challenge of recruiting patients in the late disease stages, which is often a limitation in descriptive studies (78), and showing the results obtained by all three methods of WHODAS 2.0 scoring (simple, complex and dichotomic) (82).

This study has some limitations. The main limitation is its cross-sectional design, which restricts the analysis to data collected at a single point in time. To address this limitation, we analyzed the impact of disease severity by comparing paired groups in terms of main demographic and clinical aspects. Further, longitudinal studies will be needed to investigate the long-term decline in functionality based on WHODAS 2.0.

Another limitation is that none of the participants were in stage 5 of the H&Y scale. Further studies should include patients in stage 5 to provide a comprehensive understanding of the study’s findings.

### Conclusion

The WHODAS 2.0 was able to detect a progressive decline in functionality associated with increased disease severity, indicating that some dimensions of functionality, such as mobility, independence of ADL, and self-care, are impacted earlier than others. By using the WHODAS 2.0, we were able to identify limitations not captured by traditional symptom-based scales such as the MDS-UPDRS, including difficulties in household activities, social participation, interpersonal relationships, and work-related tasks.

Incorporating WHODAS into routine clinical assessments and research can offer an innovative perspective on PD care. It enables health professionals to move beyond a symptom-focused approach towards person-centered management strategies that address functional limitations, social participation, and overall quality of life. This expanded evaluation is essential for designing targeted pharmacological and non-pharmacological interventions that meet the real-world needs of people living with PD and promote their autonomy and wellbeing across disease stages

Furthermore, the WHODAS 2.0, as a transcultural instrument for assessing functionality across diverse health conditions, is important not only for evaluating key health outcomes in PD interventions but also for facilitating meaningful comparisons with other populations affected by the same disease, as well as with people suffering from different health conditions.

## Materials and Methods

### 2.1 Study Design and Participants

A cross-sectional study was conducted according to the Strengthening the Reporting of Observational Studies in Epidemiology (STROBE) statement, based on a convenience sample of 352 people with PD. The eligibility criteria were (a) confirmed diagnosis of idiopathic PD according to the diagnostic criteria of the UK Parkinson’s Disease Society Brain Bank (83), (b) in stages 1-4 according to H&Y (19), (c) aged between 35-85 years of age, (d) using dopaminergic medication, (e) who had access to the internet for remote interviews and agreed to participate in the study. The non-eligibility criteria were (a) the presence of neurological disorders other than PD, (b) the presence of another chronic debilitating disease such as osteoarthritis, chronic obstructive pulmonary disease, or cancer, (c) diagnosis of major depression, and (d) significant cognitive, speech, and hearing disorders that could impair the remote interview feasibility.

The ability of participants to respond accurately to questions regarding personal and socioeconomic information in the first section of the study questionnaire was used as clinical evidence of their minimal cognitive capacity to understand the study’s objectives and the content of the questions. Their responses were confirmed by their family or caregivers

The participants were divided into four groups according to H&Y stages (1–4).

This article was produced as part of the activities of FAPESP Research, Innovation and Dissemination Center for Neuromathematics (grant #<u>2013/ 07699-0</u>, S.Paulo Research Foundation). It was approved by the Ethics Committee of the Department of Physiotherapy, Speech Therapy, and Occupational Therapy at the University of São Paulo Medical School, São Paulo, Brazil (#CAAE 19504619.5.0000.0065) and conducted in accordance with the Helsinki Declaration

### 2.2. Recruitment

Participants were recruited consecutively from the contacts of the AMPARO network (www.amparo.numec.prp.usp.br) between June 2022 and December 2023 by a non-probability sampling method. To ensure group matching by H&Y stages, participants’ age, sex, schooling, and SEC were controlled.

The clinical information was obtained from the healthcare system records where patients receive care for PD. The records were no older than six months to ensure that the information accurately reflected the participants’ current condition and to avoid outdated information. This information was subsequently verified with participants and their families. Details of dopaminergic treatment, including drug type and daily dose, were recorded to calculate the levodopa equivalent daily dose (LEDD) (84).

### 2.3. Study Procedures

The participants were requested to specify their preferred day and time for the remote interview, considering the on-period of dopaminergic medication (40-120 minutes post the last intake). Additionally, the participants could decide whether to have family assistance during the interview. The researchers applied the questionnaire that included: (1) general information; (2) information related to PD, including medication schedule for LEDD calculation (54); (3) socioeconomic condition (SEC); (4) global cognitive capacity by Telephone-Montreal Cognitive Assessment (T-MoCA); (5) self-perception of functionality by WHODAS 2.0, (6) self-perception of non-motor and moto aspects of daily life experiences of PD by Movement Disorder Society – Unified Parkinson’s Disease Rating Scale - Part I (MDS-UPDRS I) and Part II (MDS-UPDRS II) respectively; (7) depressive symptoms by Beck Depression Inventory (BDI). All interviews were conducted by researchers trained in qualitative and quantitative interview methods and familiar with the tools and the study objectives.

### 2.4 Instruments SEC

The SEC was evaluated by the Socioeconomic Stratum (SES), a socioeconomic classification standard based on households, family education, and income (85). The score ranged from zero to 100. Higher scores indicate better SEC conditions.

#### T-MoCA

The T-MoCA is an adapted version of the MoCA test administered by phone. It contains only the items that do not require pencil and paper or visual stimuli, i.e., nomination, memory, attention, language, abstraction, delayed recall, and orientation. The score ranged from 0 to 22 (86). T-MoCA effectively detected cognitive impairment with remote administration, showing a strong correlation with MoCA (87).

#### WHODAS 2.0

It is a practical, generic assessment instrument that can measure health and disability at the population level or clinical practice based on the difficulties presented in the last 30 days. There are several different versions of WHODAS 2.0, which differ in length and intended mode of administration. This study used the full version of 36 items through an interview.

WHODAS 2.0 captures the level of functioning in six domains of life: Domain 1: Cognition – Evaluates communication and cognitive functions; specific areas assessed include attention, memory, problem-solving, learning, and verbal expression (questions D1.1 - D1.5); Domain 2: Mobility – Assesses activities related to physical movement, such as standing, navigating within the home, leaving the home, and walking long distances (questions D2.1 - D2.5); Domain 3: Self-Care – Evaluates abilities related to personal hygiene, dressing, eating, and managing alone (questions D3.1 - D3.4); Domain 4: Getting Along – Assesses interactions with others and challenges that may arise in social situations due to health conditions; this includes relationships with close acquaintances (e.g., spouse, family members, friends) as well as interactions with strangers (questions D4.1 - D4.5); Domain 5: Activities Daily Living – Assesses difficulties in performing daily activities, including household tasks, leisure activities, employment, and education subdomain Household (questions D5.1 - D5.4) and subdomain Work (questions D5.5 - D5.10); Domain 6: Participation – Assesses social dimensions, such as community activities; barriers and hindrances in the world around the respondent; and problems with other issues, such as maintaining personal dignity (questions D6.1 - D6-8). The questions do not necessarily and solely refer to the ICF participation component as such but also include various contextual (personal and environmental) factors affected by the health condition of the respondent (33).

For all six domains, WHODAS 2.0 provides a profile and a summary measure of functioning and disability that is reliable and applicable by remote interview (32, 88), across cultures in all adult populations (33, 89). Each question is scored between 1 and 5. The higher the score, the worse the functionality of people.

The present study adopted “simple scoring,” “complex scoring,” and “dichotomous scoring.” In the simple scoring, the scores assigned to (1) “none”, (2) “mild”, (3) “moderate”, (4) “severe”, and (5) “extreme/cannot “ – for each question were summed. The simple sum of the scores of the items across all domains constitutes a statistic sufficient to describe the degree of functional limitations (90) and has high internal consistency (91).

The complex scoring considers multiple difficulty levels for each WHODAS 2.0 item. This type of scoring for WHODAS 2.0 allows for more fine-grained analyses that use the complete information of the response categories for comparative analysis across populations or subpopulations. It takes the coding for each item response as “none”, “mild”, “moderate”, “severe” and “extreme” separately. Then, it uses a computer to determine the summary score by differentially weighing the items and the severity levels. The scoring has three steps: (1) summing of recoded item scores within each domain; (2) summing of all six domain scores; and (3) converting the summary score into a metric ranging from 0 to 100 (where 0 = no disability; 100 = full disability). (33, 82).

Finally, we employed a dichotomous (yes/no) scoring to identify the most affected activities regardless of domain and PD stages. This scale allows respondents to indicate whether they experience no difficulty in a specific activity among the 36 examined by WHODAS (no) or if they have some difficulty (yes). The response options for “mild,” “moderate,” “severe,” and “extreme/cannot” were consolidated into a single affirmative response (33).

According to the WHODAS 2.0 guidelines, if a participant was not working, the total score was calculated using the responses from the other 32 questions. In cases where there were missing responses—no more than one per domain and no more than two in total—the median score from the other questions within the same category was used to fill in the missing data (33).

#### MDS-UPDRS

It is a tool to measure the severity and progression of PD based on the difficulties presented in the last seven days. It consists of four parts: Part I (non-motor experiences of daily living), Part II (motor experiences of daily living), Part III (motor examination) and Part IV (motor complications). In this study, only Part I and Part II were used. The Part I includes 13 questions that assess the impact of non-motor symptoms on ADL. It is correlated strongly with other validated scales for non-motor alterations in PD (75). Part II contains 13 questions that assess motor impact on ADL. It is effective for evaluating disability in PD (76). Each question is scored between 0 and 4. The higher the score, the worse the severity and progression of PD. Part I and II scores ranged from 0 to 52 (92).

#### BDI

The BDI is a widely utilized self-report questionnaire designed to assess the severity of depressive symptoms in individuals aged 13 and above. The BDI consists of 21 items, each representing a symptom of depression such as sadness, guilt, or fatigue. Respondents rate the severity of each symptom over the past two weeks. The score ranged from 0 to 63, with higher scores indicating more severe symptomatology. Scores between 0-9 indicate that the individual is not depressed, 10-18 indicate mild to moderate depression, 19-29 indicate moderate to severe depression, and 30-63 indicate severe depression. (93)

### 2.5. Statistical Analysis

Descriptive statistics of demographic, clinical, and therapeutic data for each H&Y stage 1-4 were provided. The normal distribution of the samples was assessed by the Kolmogorov-Shapiro test. Categorical variables are reported in count and percentage, and variables in means and standard deviations (SD) or medians and interquartile ranges (IQR) according to their distribution. A descriptive analysis of the WHODAS 2.0 total score and seven subscores (the results related to home and work activities from Domain 5 were analyzed separately) was performed.

For variables with normal distribution (age), the group differences in the H&Y stage were tested by One-Way ANOVA. Tukey post-test was applied to pair-to-pair comparisons when statistically significant differences were observed.

For the non-parametric variables, group differences (H&Y stages) were tested by Kruskal-Wallis. When statistically significant differences were observed, multiple comparisons of the average ranks for each pair of groups were applied; normal z-values were computed for each comparison, and Dunn post-hoc (corrected for the number of comparisons) was used for a two-sided test of significance.

Nonparametric Spearman’s rank correlation coefficients were calculated to analyze the association between variables (coefficients higher than 0.59 were considered “high”).

A significance level of p < 0.05 was used to determine the statistical significance of the findings. All statistical analyses were performed using Statistica Version 13 (TIBCO Software Inc. USA).

## Data availability

All data generated or analyzed during this study are included in this published article [and its supplementary information files].

## Supplementary Information

**Supplementary Table 1:**
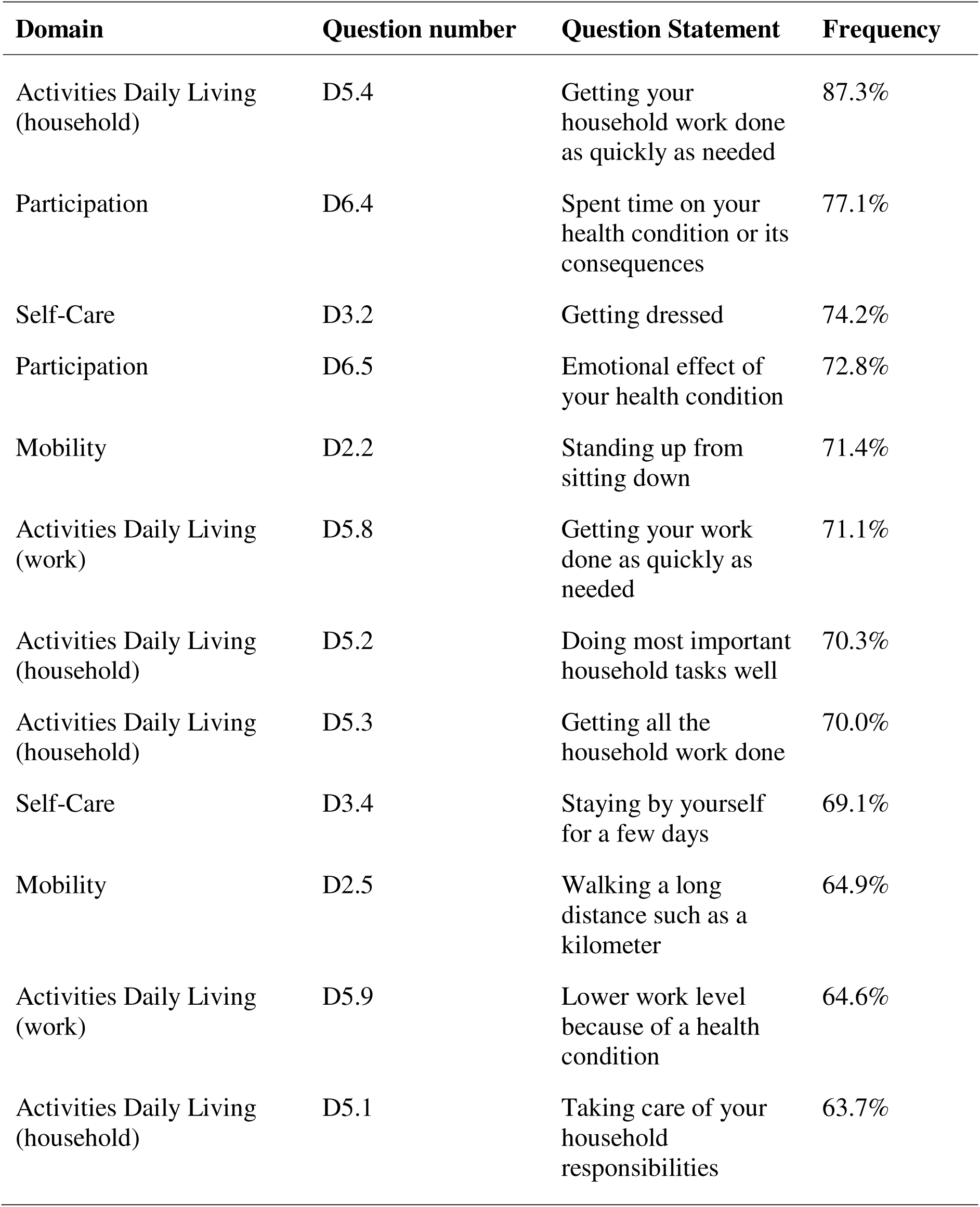
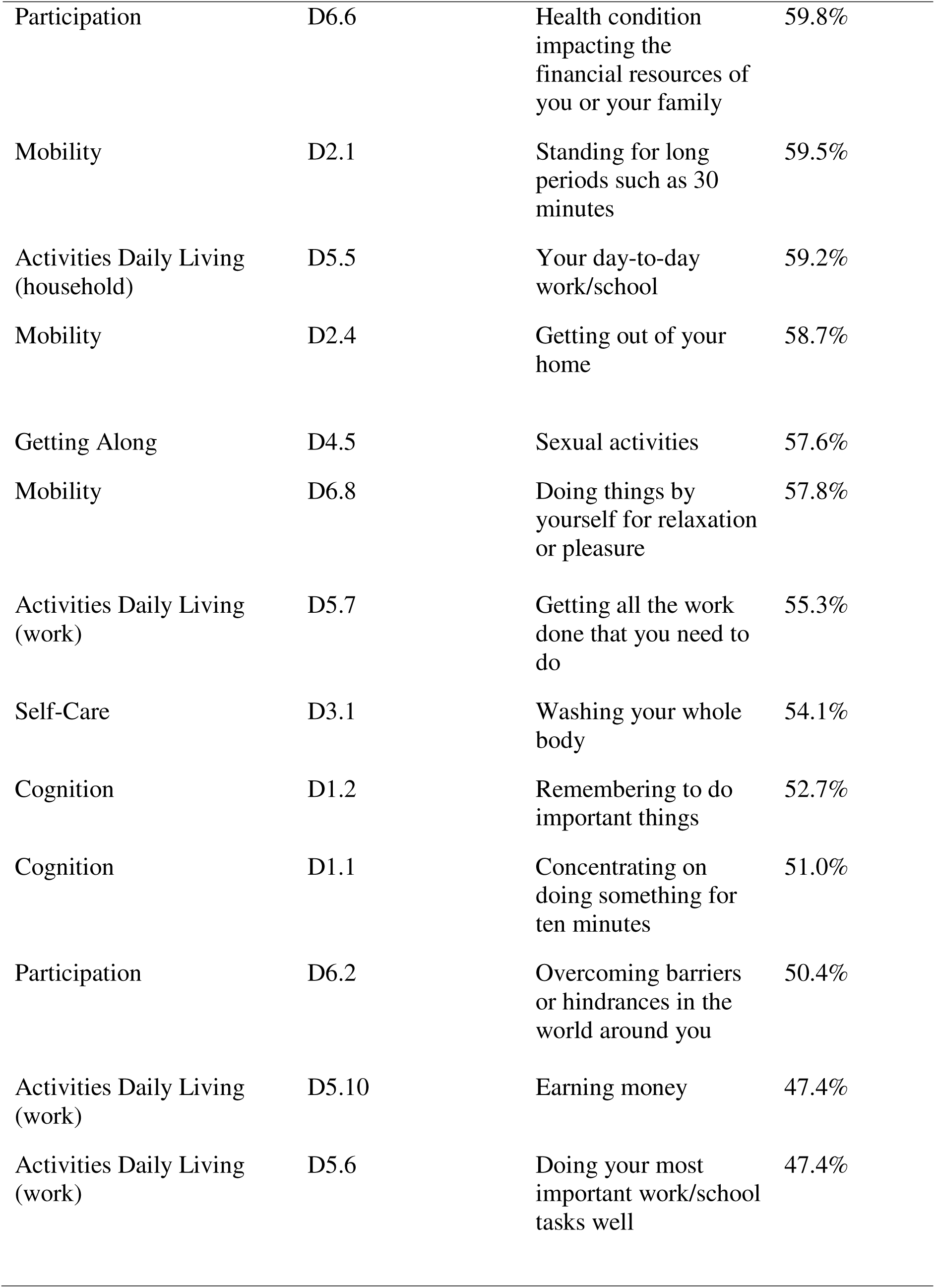
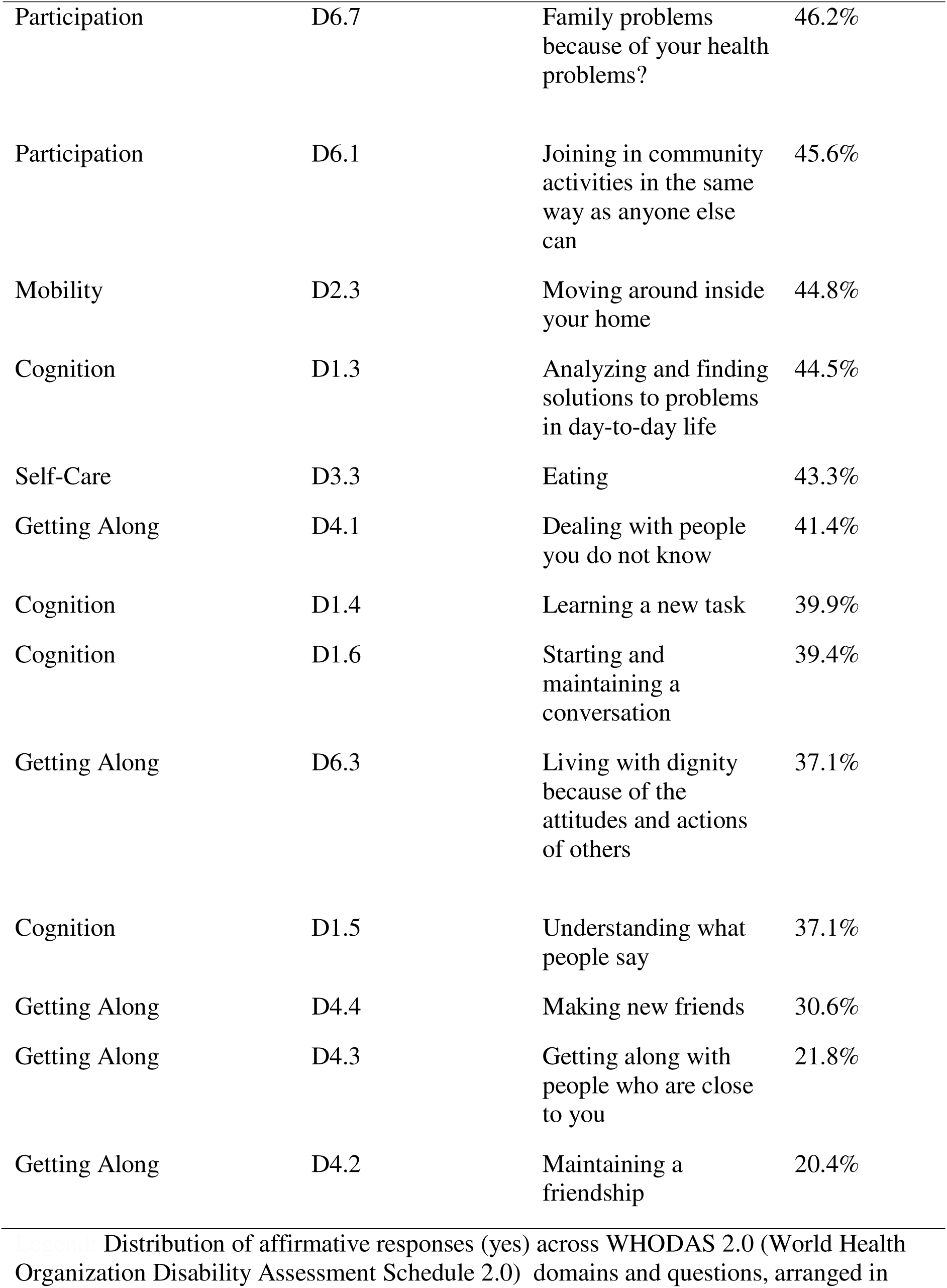
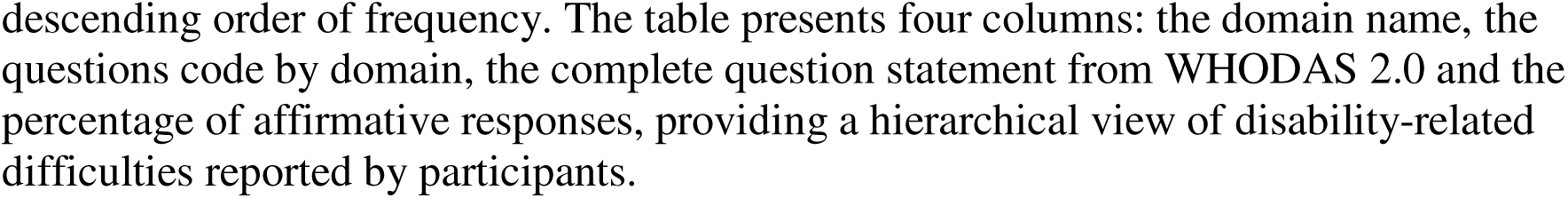
Frequency of disability-related difficulties for each WHODAS 2.0 question.

**Supplementary Table 2:** Mean scores of functioning and disability across WHODAS 2.0 domains.

| Domain | Description | Mean Score |
| --- | --- | --- |
| Domain 1 - Cognition | Communication and thinking activities | 1.75 |
| Domain 2 - Mobility | Moving and getting around | 2.16 |
| Domain 3 - Self-Care | Hygiene, dressing, eating, staying alone | 1.99 |
| Domain 4 - Getting Along | Interacting with other people | 1.66 |
| Domain 5h - Activities Daily Living (household) | Domestic responsibilities | 2.31 |
| Domain 5w - Activities Daily Living (work) | Work/school activities | 1.40 |
| Domain 6 - Participation | Joining in community activities | 2.16 |

**Supplementary Table 3:**
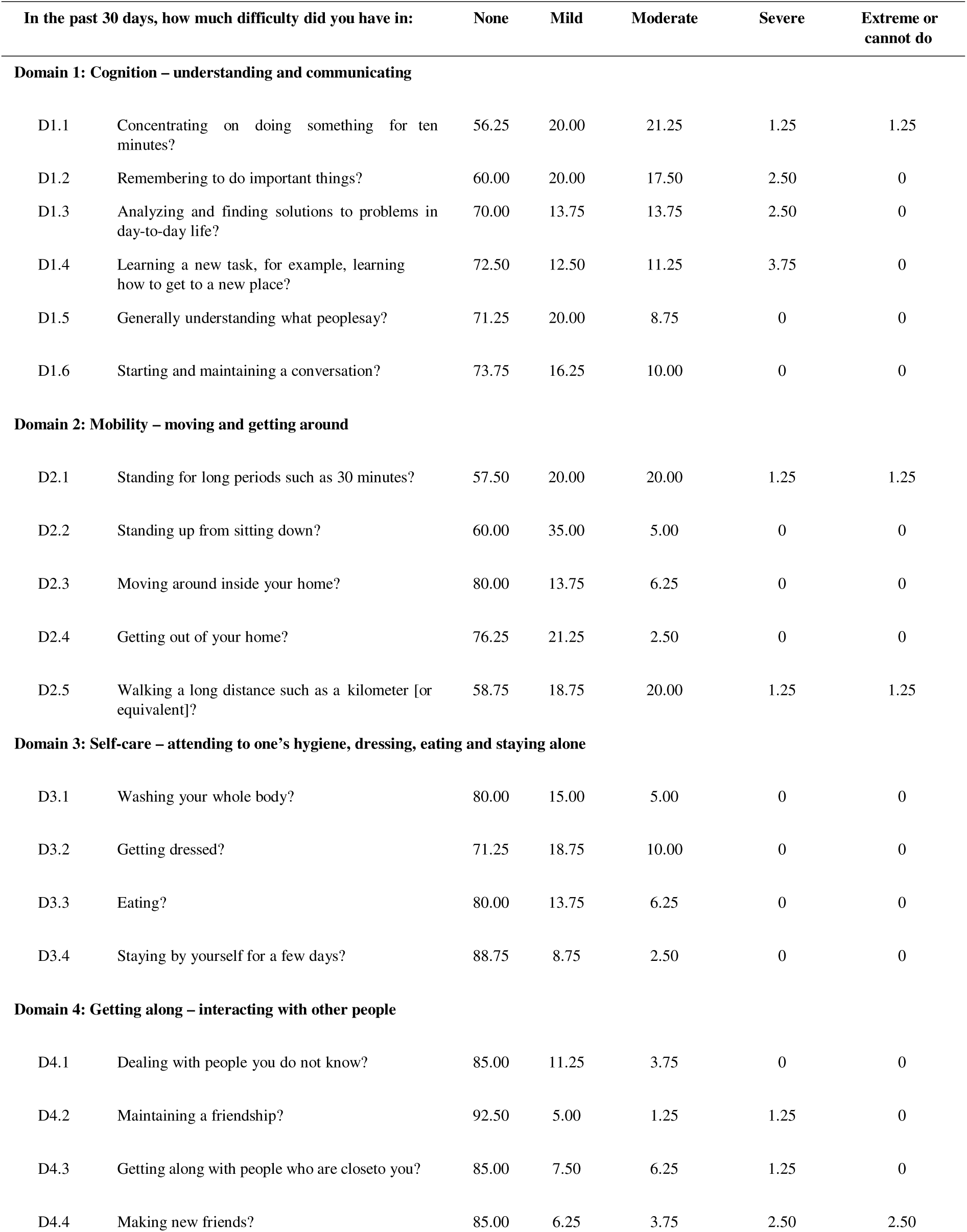
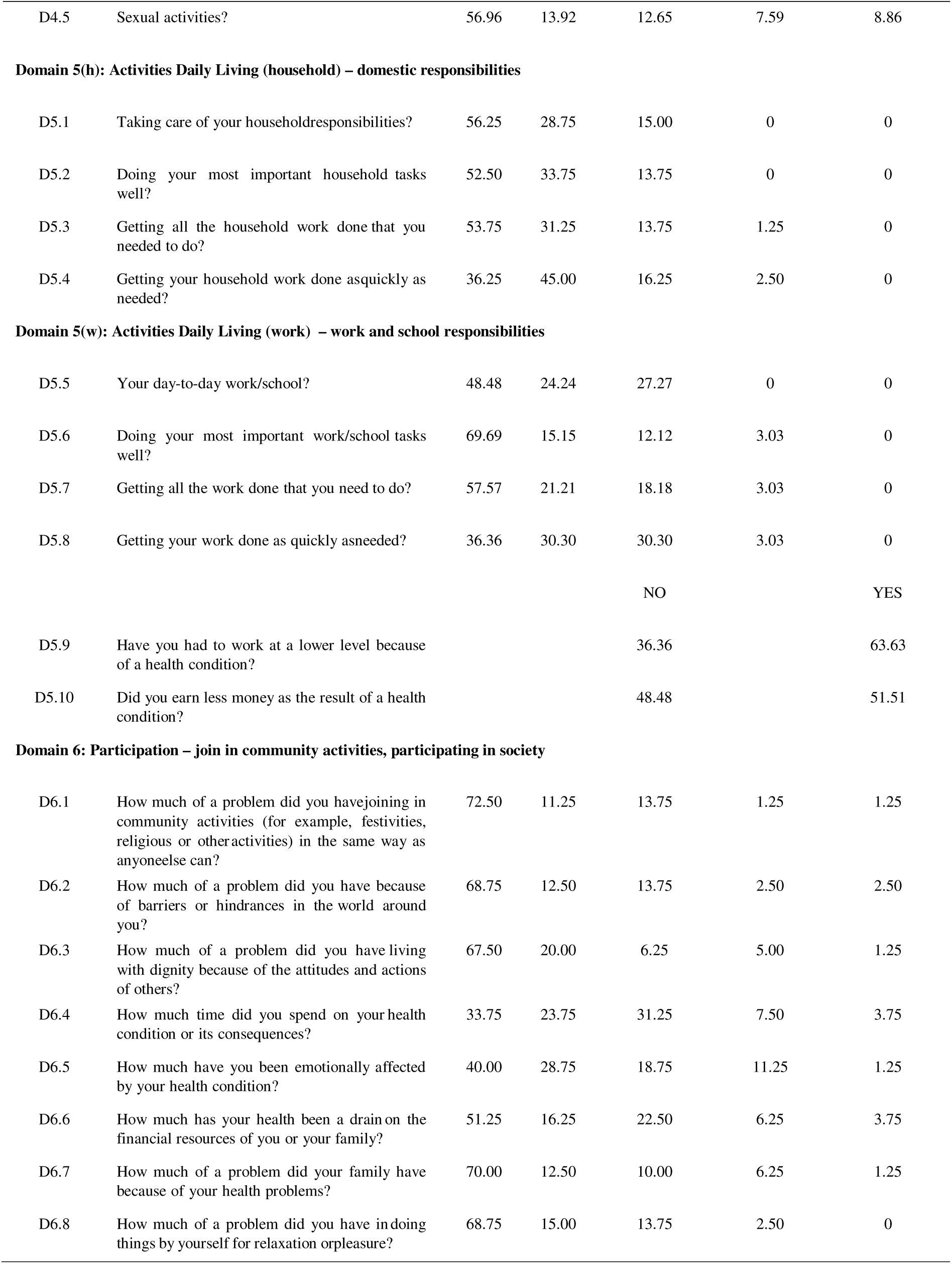
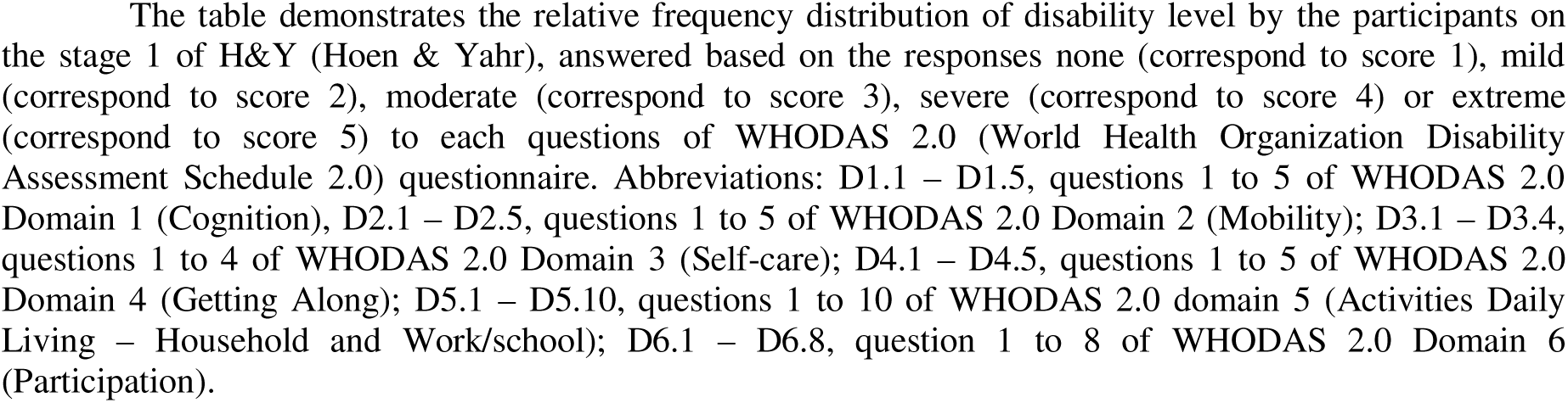
Relative frequency distribution of disability level in participants in stage 1 of H&Y according to WHODAS 2.0 domains.

**Supplementary Table 4:**
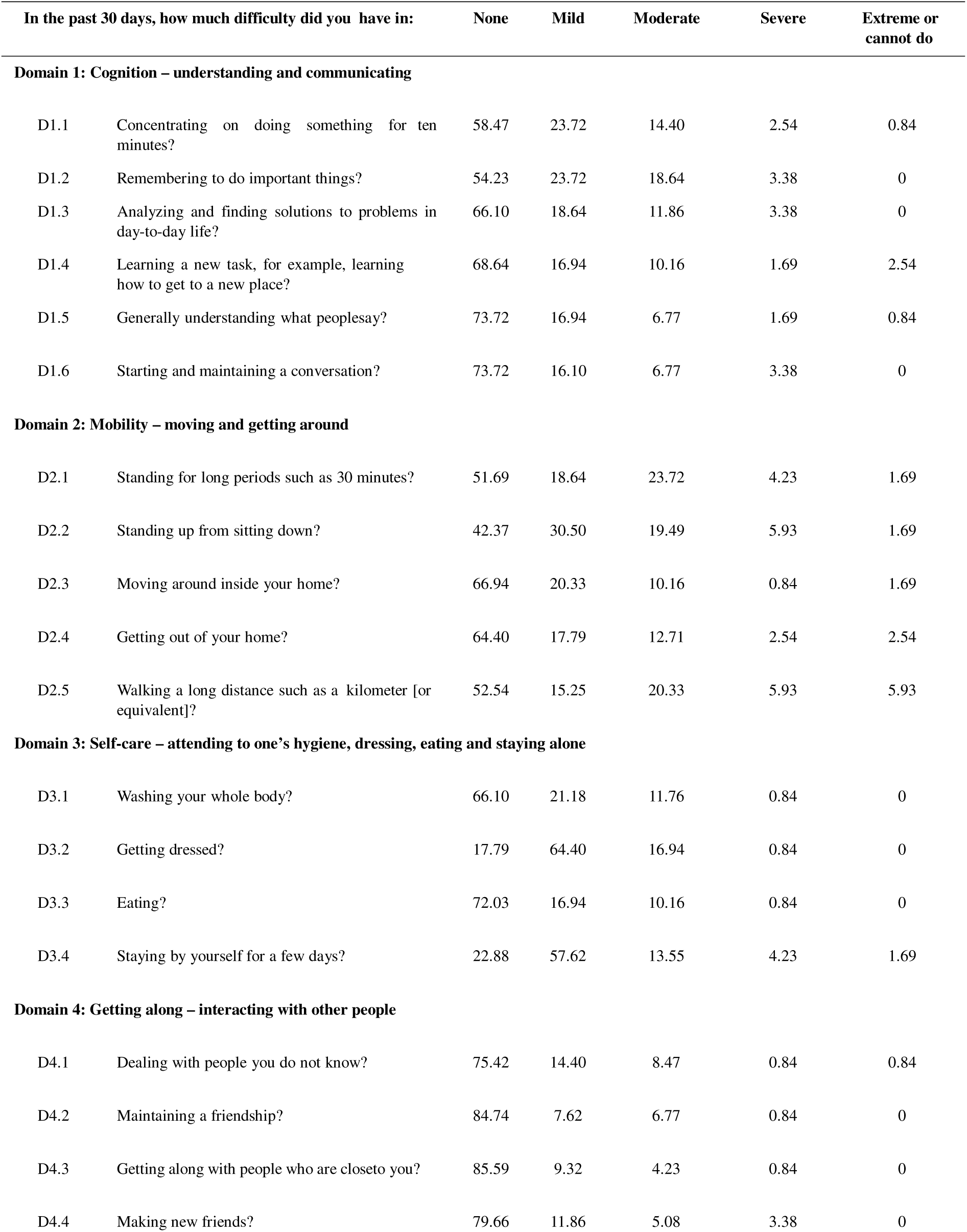
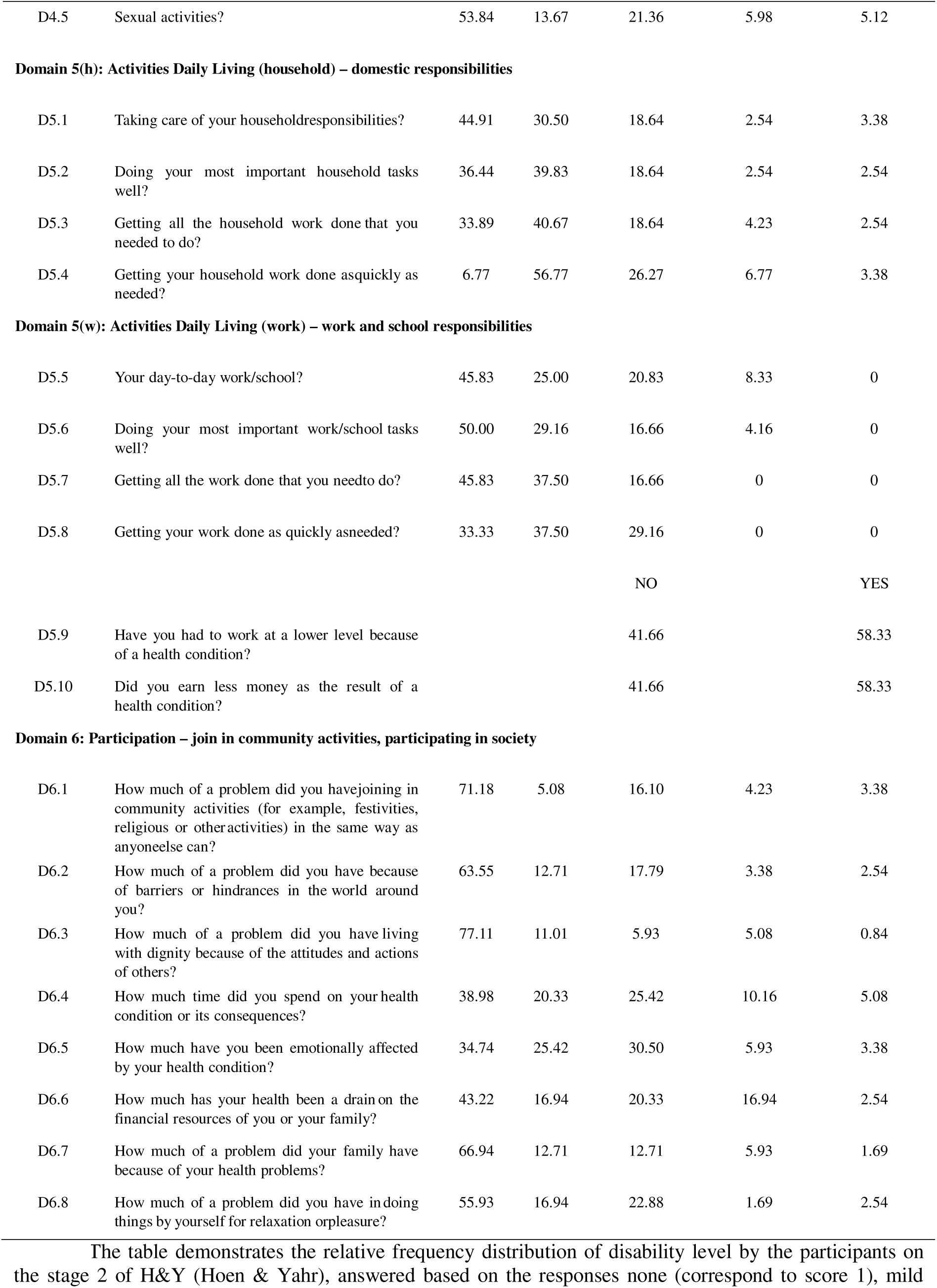
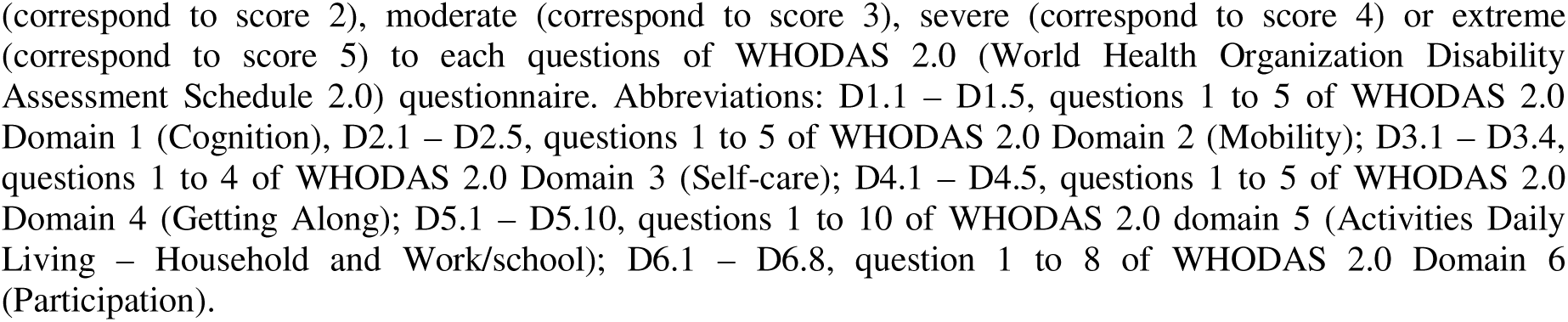
Relative frequency distribution of disability level in participants in stage 2 of H&Y according to WHODAS 2.0 domains.

**Supplementary Table 5:**
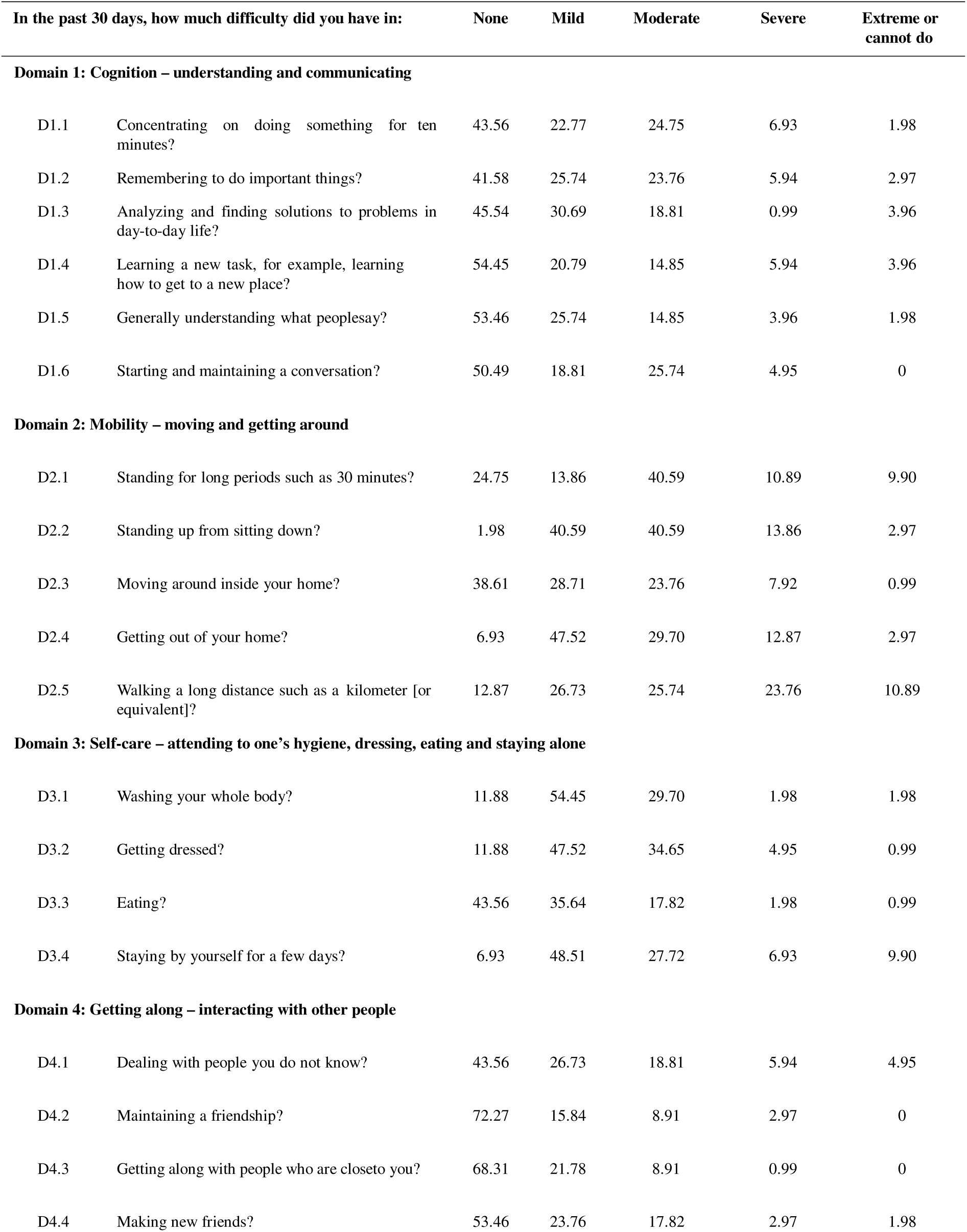
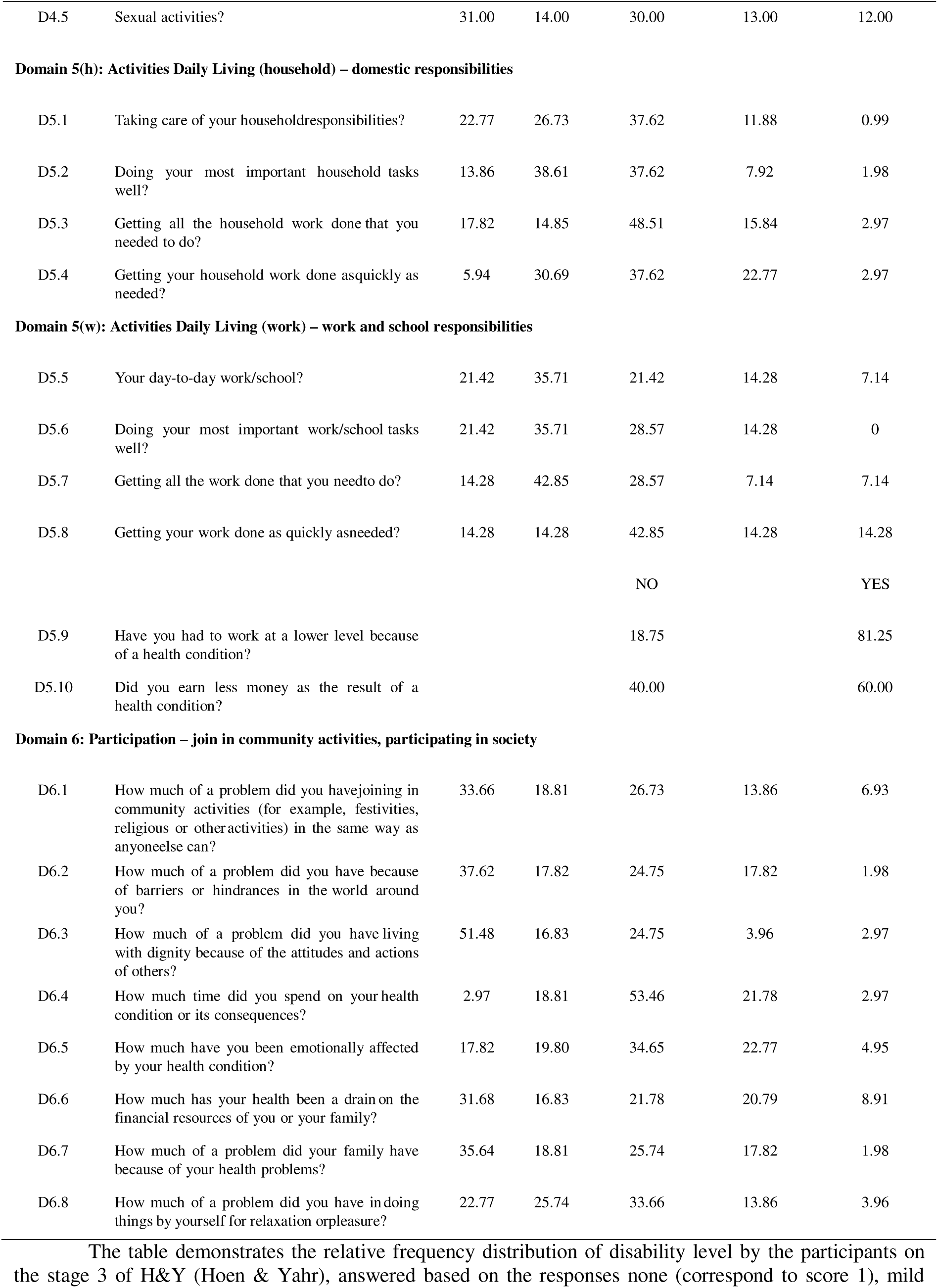
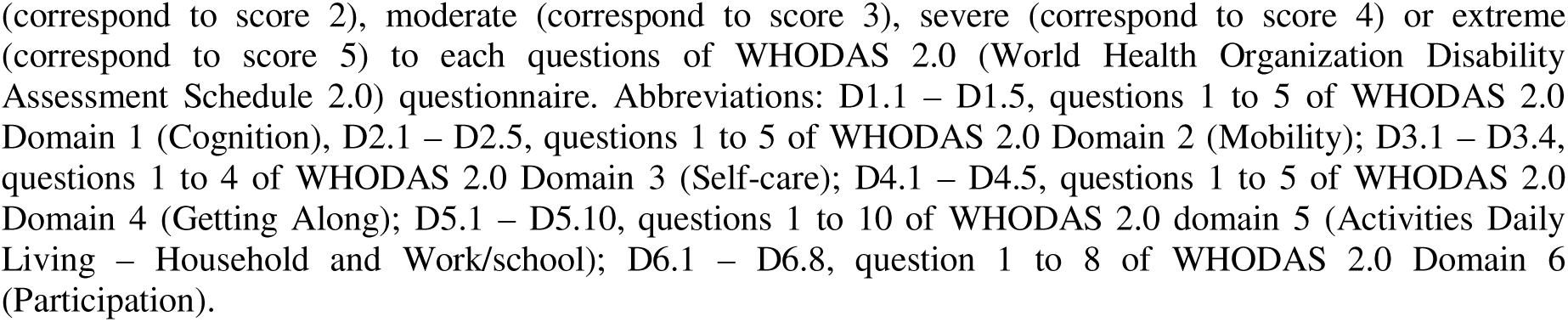
Relative frequency distribution of disability level in participants in stage 3 of H&Y according to WHODAS 2.0 domains.

**Supplementary Table 6:**
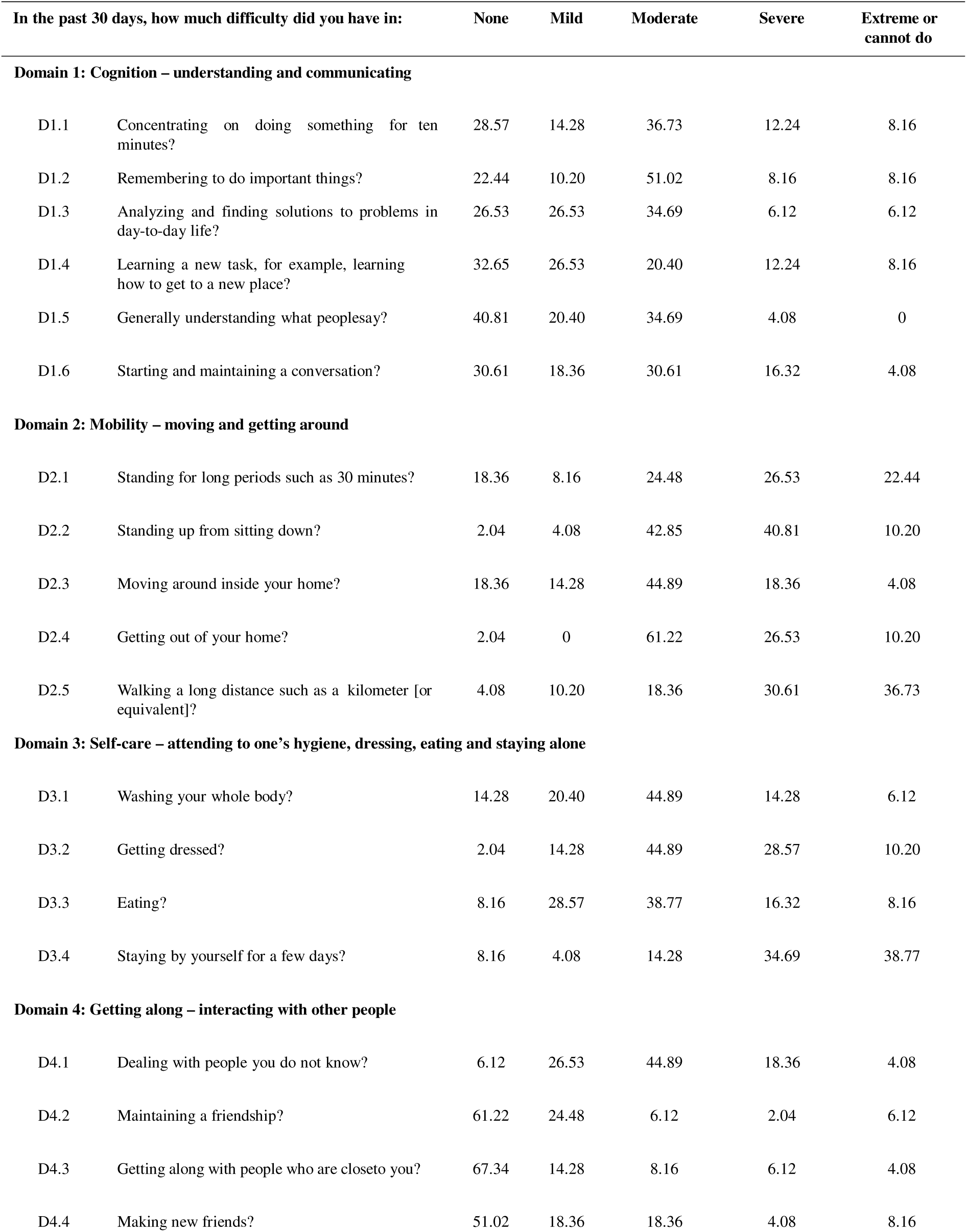
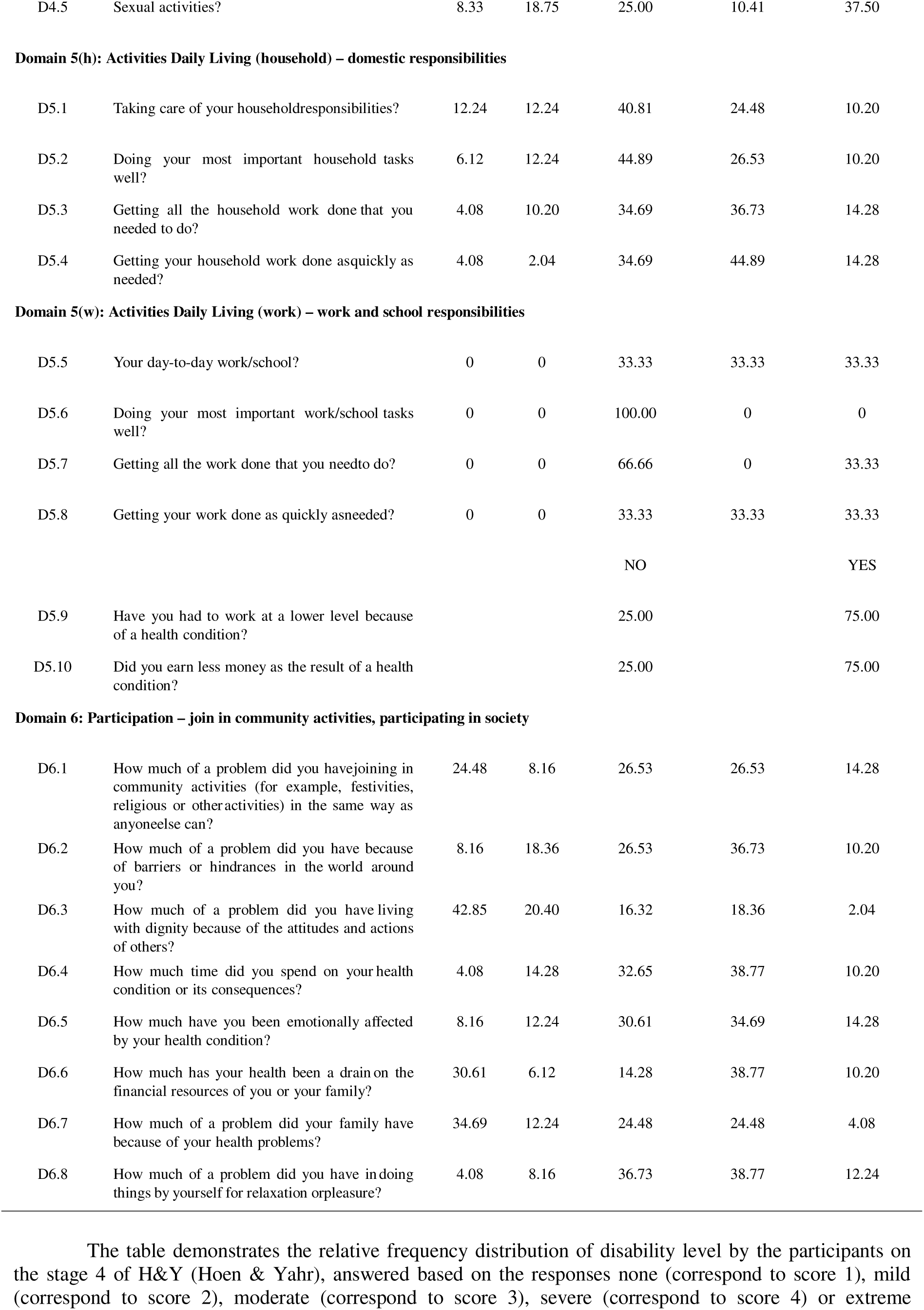
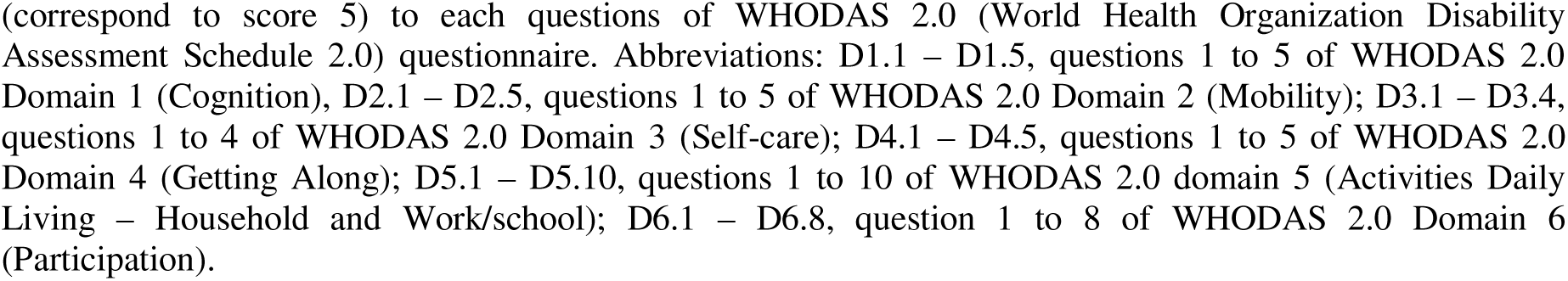
Relative frequency distribution of disability level in participants in stage 4 of H&Y according to WHODAS 2.0 domains.

